# High-Intensity Interval Training Reduces Transcriptomic Age: A Randomized Controlled Trial

**DOI:** 10.1101/2022.12.22.22283853

**Authors:** Trevor Lohman, Gurinder Bains, Steve Cole, Lida Gharibvand, Lee Berk, Everett Lohman

## Abstract

**Background:** The relationship between exercise and lifespan is well documented, but little is known about the effects of specific exercise protocols on modern measures of biological age. Transcriptomic age predictors provide an opportunity to test the effects of high intensity interval training (HIIT) on biological age utilizing whole-genome expression data.

**Methods:** A single-site, single-blinded, randomized controlled clinical trial design was utilized. Thirty sedentary participants (aged 40 to 65) were assigned to either a HIIT group or a no-exercise control group. After collecting baseline measures, HIIT participants performed three 10×1 HIIT sessions per week for 4 weeks. Each session lasted 23 minutes, and total exercise duration was 276 minutes over the course of the 1-month exercise protocol. Transcriptomic age, PSS-10 score, PSQI score, PHQ-9 score, and various measures of body composition were all measured at baseline and again following the conclusion of exercise/control protocols.

**Results:** Transcriptomic age reduction of 3.59 years was observed in the exercise group while a 3.28-year increase was observed in the control group. PHQ-9, PSQI, BMI, body fat mass, and visceral fat measures were all improved in the exercise group. A hypothesis-generation gene expression analysis suggested exercise may modify autophagy, mTOR, AMPK, IP3K, neurotrophin signaling, insulin signaling, and other age-related pathways.

**Conclusion:** A low dose of HIIT can reduce an RNA-based measure of biological age in sedentary males and females between the ages of 40 and 65. Other changes to gene expression were relatively modest, which may indicate a focal effect of exercise on age-related biological processes.

## Introduction

The beneficial effects of exercise on healthspan and lifespan are among the most well documented scientific findings in health science research (Aune et al., 2021; Han et al., 2022; Myers et al., 2002; Northey, Cherbuin, Pumpa, Smee, & Rattray, 2018). Despite this, there are relatively few trials investigating the effects of exercise on gene regulatory mechanisms of healthspan and lifespan. Of those that have been performed, most examine the effects of a single bout of exercise on gene expression, rather than repeated bouts (Amar et al., 2021).

Given that many beneficial effects of exercise require repeated bouts over time to manifest, this represents an opportunity for discovery. Consider for example the inappropriate conclusions that could be drawn when studying the effects of a single bout of exercise on muscle hypertrophy, strength, or inflammation. The beneficial effects of exercise on biological aging are likely most apparent when studied over time.

The central theme of molecular biology holds that a cell’s function and status are dictated by the specific sets of genes undergoing transcription at any given time, and to what degree these processes are occurring (O’Brien, Costin, & Miles, 2012). Genome-wide expression analyses allow us to take a snapshot of those processes, capturing a gene expression profile at the time of blood draw. A comparison of gene expression profiles before and after an intervention provides the means to identify patterns of differentially expressed genes.

As high throughput RNA sequencing becomes more commonplace, gene expression-based predictive models have emerged. Some of these models are designed to predict biological age (Meyer & Schumacher, 2021; Peters et al., 2015; Ren & Kuan, 2020), or more specifically, transcriptomic age (TA). These models are easily accessible and comprehensive molecular surveys of biological processes that collectively contribute to healthspan and lifespan. It is this type of biological age predictor, a “transcriptomic clock” that is used in the trial described here.

The biological age prediction field is diverse and rapidly evolving, with models composed of various inputs (Cesari, Gambassi, Abellan van Kan, & Vellas, 2014; Jylhävä, Pedersen, & Hägg, 2017; Levine et al., 2018; Lohman, Bains, Berk, & Lohman, 2021; Lu et al., 2019) and predictive capabilities (Li et al., 2020; McCrory et al., 2020). The discrepancy between a participant’s actual age and their predicted age is often of particular interest (Fahy et al., 2019; Fiorito et al., 2021). This measure, called age acceleration (biological age minus chronological age), can take a positive or negative value. Positive values are considered hazardous and indicative of an increased aging rate, while negative values are considered beneficial and evidence of a slowed aging rate. Any intervention that reverses age acceleration could therefore be considered beneficial and potentially health protective.

The effect of exercise on various biological age predictors is inconsistent. Most experimental studies that examine the relationship between exercise and biological age use telomere length as their primary biomarker of aging. These results are mixed, with positive relationships, U-shaped relationships, and no relationship all being reported (Sellami, Bragazzi, Prince, Denham, & Elrayess, 2021). This could be due to any number of factors, from differences in sample characteristics to the open question of whether telomere length even has utility as a measure of biological age (Glei et al., 2016; Li et al., 2020; Svensson et al., 2014; Vaiserman & Krasnienkov, 2020; Wang, Zhan, Pedersen, Fang, & Hägg, 2018).

Fewer studies have been performed using epigenetic alteration, such as DNA methylation or histone methylation/acetylation as an outcome measure. Of those that have been performed, various types of exercise have been shown to induce widespread changes to the methylome and associated gene expression (Barrès et al., 2012; Denham, O’Brien, Marques, & Charchar, 2015; Nakajima et al., 2010).

To the authors’ knowledge only two lifestyle modification trials have utilized a next generation predictor of biological age in humans, such as an epigenetic clock (Fiorito et al., 2021; Fitzgerald et al., 2020), and no prior study has used a transcriptomic predictor of biological age (Nielsen, Bakula, & Scheibye-Knudsen, 2022).

The trial described here aims to address this by utilizing high throughput RNA sequencing to explore the effects of twelve high intensity interval training (HIIT) sessions on biological age as measured by a blood mRNA-based “transcriptomic clock” (Peters et al., 2015).

To confirm previously observed effects of HIIT on various physiological parameters (Gu, Hao, Chen, & Wu, 2022; Min, Wang, You, Fu, & Ma, 2021; Ouerghi et al., 2017; Su et al., 2019; M. Wewege, van den Berg, Ward, & Keech, 2017) we also measured changes to body mass index (BMI), body fat mass (BFM) and visceral fat area, as well as measures of psychological stress, depression, and sleep quality.

## Methods

A randomized controlled trial design was used to investigate the effects of HIIT on the following dependent variables: 10-item Perceived Stress Scale (PSS-10) (Lee, 2012), Pittsburgh Sleep Quality Index (PSQI) (Zhang et al., 2020), Patient Health Questionnaire 9-item depression module (PHQ-9) (Kroenke, Spitzer, & Williams, 2001; Levis et al., 2020), body mass index (BMI), body fat mass, visceral fat area, skeletal muscle mass, waist-to-hip ratio, blood pressure, resting heart rate, and whole-genome RNA expression. The transcriptomic age prediction (TRAP) tool (Peters et al., 2015) was used to assess transcriptomic age and transcriptomic age acceleration (TAaccel = TA – chronological age) using the RNA AGE Calc Shiny App (Ren & Kuan, 2020). The TRAP biological age prediction model was trained to predict chronological age in a meta-analysis of 14,983 individuals and is based on 11,908 input gene expression levels (Peters et al., 2015).

Trial participants were recruited from local communities surrounding the Loma Linda University campus via recruitment flyers, approved social media, and word of mouth. The Loma Linda University Institutional Review Board approved the study on 11/18/2021 (IRB# 5210437, clinicaltrials.gov trial registration ID: NCT05156918). Males and females between the ages of 40 and 65 who self-identified as non-exercisers, were categorized as low activity using the International Physical Activity Questionnaire (IPAQ) (Hagströmer, Oja, & Sjöström, 2006), had no significant change to activity levels within the past 30 days, were not pregnant, had no prior or current history of any condition that would make exercise unsafe, and were not currently taking antibiotics, glucocorticoids, anticoagulants, narcotics, antiepileptics, antipsychotics, or hypoglycemic agents were eligible for participation.

Study participants were instructed to avoid modifying their usual physical activity level or diet for the duration of the four-week study protocols, except for the additional HIIT assigned to exercise group. All participants maintained a compliance log, comprised of two questions weekly. For the control group: Have you performed more than your usual amount of physical activity this week? Secondly, have you made any significant changes to your diet this week? For the exercise group: Excluding the exercise assigned to you in this study, have you performed more than your usual amount of physical activity this week? Secondly, have you made any significant changes to your diet this week?

All participants arrived at the laboratory between the hours of 8am and 11am, and baseline measures were obtained. Body composition measurements were obtained using the InBody 770 body composition and body water analyzer (InBody USA, USA), surveys were completed in a private room, and a single vial of blood was collected by a certified phlebotomist from the antecubital vein into a PAXgene® Blood RNA Tube, PLH 16×100 2.5 PLBLCE CLR (Becton Dickinson, USA)

Following the completion of Day-1 data collection, exercise group participants returned the following day to begin the HIIT protocol which took place at the Loma Linda University Physical Fitness Laboratory. The authors chose a routinely studied 10×1 HIIT protocol that has been determined as safe and effective in various groups, including sedentary individuals (Ito, 2019; Little, Safdar, Wilkin, Tarnopolsky, & Gibala, 2010; Rozenek, Salassi, Pinto, & Fleming, 2016; M. A. Wewege, Ahn, Yu, Liou, & Keech, 2018). The protocol consists of a 2-minute warm up and cool down, with 10, 1-minute high intensity exercise intervals at 77-93% of the participants predicted maximum heart rate (Committee; Riebe, Ehrman, Liguori, & Magal) determined using Karvonen’s formula (Camarda et al., 2008), followed by 1-minute self-selected intensity rest periods. The total exercise session lasted 23 minutes, of which 10 minutes was high intensity exercise and 13 minutes was warm-up/rest/cool down periods.

Participants rotated between three exercise machines (randomly assigned rotation order at outset): A Concept2 rowing ergometer, Concept2 bicycle ergometer, and a Noraxon PhysTread Pressure treadmill. Participants used a different machine each day so that they used each of the three exercise machines once per week.

Following the conclusion of the 4-week control and exercise protocols, all participants returned for results collection. Exercise group data was collected approximately 48 hours after their last HIIT session. Blood samples were stored at -79 degrees Celsius until RNA extraction (Qiagen RNeasy), quality assurance assays, mRNA sequencing, and related statistical analyses of differential gene expression and interpretive bioinformatics were performed by the UCLA Social Genomics Core Laboratory. Transcriptional profiling utilized a high-efficiency mRNA targeted reverse transcription and cDNA library synthesis system (QuantSeq 3’ FWD; Lexogen Inc.) with cDNA libraries sequenced on in Illumina NovaSeq system by Lexogen Services GmbH. Assays targeted 5 million sequencing reads per sample (achieved median = 7.1 million), each of which was mapped to the GRCh38 reference human transcriptome using the STAR aligner (median 99.7% mapping rate) and quantified as gene transcripts per million total mapped reads, with values floored at 1 transcript per million to suppress spurious low-range variability, and log2-transformed to stabilize variance. One follow-up sample yielded insufficient sequencing reads for valid analysis (< 1 million reads), and that sample and its paired pre-intervention baseline sample were excluded from all subsequent analyses. These data served as input into the RNA AGE Calc Shiny App for computation of the TRAP RNA age score. RNA AGE Calc Shiny App inputs were as follows: Tissue type: Blood, type of gene expression data: Count, samples used when building the calculator: All samples, gene ID type: Ensembl ID, signature: Peters.

A secondary analysis of differentially expressed genes (DEGs) was performed using two sets of cut off criteria. First, genes which displayed a group x time interaction expression fold change greater than 1.5 or less than .5 were selected for analysis. Also, an exploratory/hypothesis-generation analysis was performed using more liberal fold change values, greater than 1.2 or less than .8. Functional enrichment and pathway analyses were performed using Advaita Bio’s iPathway Guide (Supplementary File 2).

### Data Analysis

Mean ± SD was computed for quantitative variables and frequency (percentage) for categorical variables. Normality of quantitative variables was assessed using Shapiro-Wilk test and box plots. Independent t-test was used for all continuous and independent variables in both groups at baseline. The Mann-Whitney U test was used to compare the same variables due to small sample and lack of normality on some variables. The dependent paired t test was used to compare pre- and post-variables in both groups. Also, Wilcoxon Signed Rank test was used to compare the pre- and post-variables due to small sample and lack of normality on some variables.

Data were analyzed using SPSS Statistics Software version 28.0 (SPSS Inc, Chicago, IL, USA). All analyses were performed at an alpha level of .05.

## Results

Of the 35 participants screened, 30 subjects satisfied the eligibility criteria, agreed to participate, were randomly assigned to the experimental group (n=15) and the control group (n=15) using computer-generated block randomization, and completed all subsequent analyses (Figure 1).

**Figure 1:**
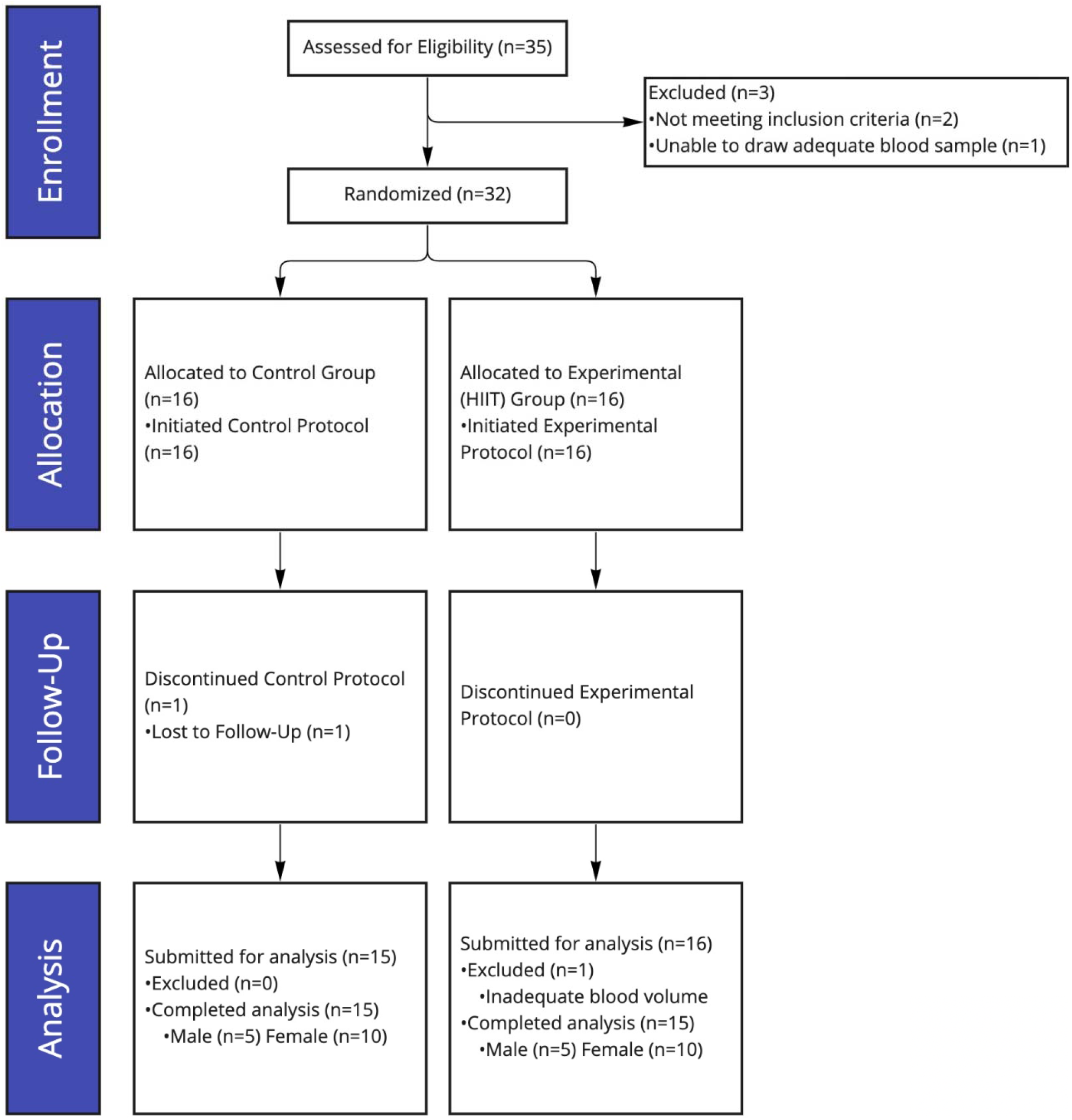
CONSORT chart diagram. 35 participants recruited, 2 excluded due to high activity level, 1 excluded due to an inability to draw blood sample. 1 control participant lost to follow up, 1 exercise participant excluded from analysis due to low blood volume in post exercise blood sample detected during RNA quality control tests. In total, 15 control participants and 15 experimental participants completed all aspects of the trial and subsequent analysis.

Baseline characteristics of participants are shown in Table 1. None of the demographic variables were significant for randomized design.

**Table 1.** Selected Characteristics of Participants at Baseline.

| Variables | Experimental<br>Frequency (%)<br>(n=15) | Control<br>Frequency (%)<br>(n=15) |
| --- | --- | --- |
| Age (years) | 51.00±7.9<sup>δ</sup> | 47.93±7.6<sup>δ</sup> |
| BMI (kg/m<sup>2</sup>) | 31.08±4.9<sup>δ</sup> | 29.59±5.4<sup>δ</sup> |
| Race/Ethnicity |  |  |
| White | 7 (46.7) | 5 (33.3) |
| Black | 2 (13.3) | 1 (6.7) |
| Hispanic | 4 (26.7) | 4 (26.7) |
| Asian | 1 (6.7) | 5 (33.3) |
| Other | 1 (6.7) | 0 (0) |
| Sex |  |  |
| Female | 10 (66.7) | 10 (66.7) |
| Male | 5 (33.3) | 5 (33.3) |
| Diabetic |  |  |
| No | 13 (86.7) | 14 (93.3) |
| Yes | 0(0) | 0 (0) |
| Pre-Diabetic | 2 (13.3) | 1 (6.7) |

<sup>δ</sup> Values are presented as mean ± SD

### Intervention Validation

There was a significant decrease in body fat mass, BMI, and visceral fat area (p= .031, .048, and .015 respectively) (Table 2), over time for the experimental group, a non-significant increase in BFM in the control group (p=.244), and a non-significant decrease in BMI and Visceral Fat Area in the control group (p=.598 and p=.062 respectively) (Table 2). No changes in body composition displayed group x time statistical significance.

**Table 2.** Effects of High Intensity Interval Training on Transcriptomic Age, PHQ-9, PSS-10, PSQI, Skeletal Muscle Mass, Body Fat Mass, and Visceral Fat Area. Between and Within Group Effects.

| Variables | Experimental (n=15) |  |  | Control (n=15) |  |  | P<sup>**</sup> |
| --- | --- | --- | --- | --- | --- | --- | --- |
|  | Pre | Post | Mean difference (P<sup>*</sup>) | pre | post | Mean difference (P<sup>*</sup>) |  |
| TA (years) | 73.4±8.2 | 69.8±7.7 | -3.59±7.72 (.043) | 67.8±9.3 | 71.1±9.2 | 3.28±8.26 (.018) | .026 |
| TAaccel (years) | 21.8±7.6 | 17.9±9.2 | -3.84±7.98 (.078) | 19.2±7.9 | 22.4±6.8 | 3.21±8.26 (.156) | .025 |
| PHQ-9 | 5.3±3.9 | 2.3±1.9 | -3.07± 3.10 (.002<sup>b</sup>) | 6.9±6.9 | 7.0±5.9 | .07± 6.15 (.964<sup>b</sup>) | .063<sup>a</sup> |
| PSS-10 | 20.1±5.3 | 19.8±4.0 | -.33±5.89 (.53<sup>b</sup>) | 21.2±3.4 | 19.7±5.2 | 1.47±4.09 (.054<sup>b</sup>) | .739<sup>a</sup> |
| PSQI | 7.0±3.9 | 5.5±3.5 | -1.53± 2.42 (.042<sup>b</sup>) | 7.6±4.7 | 7.7±4.4 | .07± 2.55 (.670<sup>b</sup>) | .158<sup>a</sup> |
| SMM (lbs) | 69.5±11.7 | 69.6±11.5 | .15± 1.39 (.676) | 63.1±14.0 | 63.5±14.0 | .39± 1.99 (.456) | .705 |
| BFM (lbs) | 74.6±22.1 | 73.1±22.2 | -1.47± 2.29 (.031<sup>b</sup>) | 66.9±22.1 | 67.0±22.7 | .17± 4.5 (.244<sup>b</sup>) | .263<sup>a</sup> |
| BMI (kg/m<sup>2</sup>) | 31.1±4.9 | 30.9±5.0 | -.23±.40 (.048) | 29.6±5.4 | 29.5±5.1 | -0.08±.57 (.598) | .513 |
| Visceral Fat Area cm<sup>2</sup> | 162.3±46.2 | 158.1±46.1 | -4.25± 5.95 (.015) | 157.0±58.3 | 154.3±58.3 | -2.66± 5.08 (.062) | .426 |

Values are presented as mean ± SD
\* p- values for the null hypothesis that there is no difference between pre and post.
\*\* p- values for the null hypothesis that there is no difference between groups.
a: Mann-Whitney U test
b: Wilcoxon Signed Rank test
Abbreviations. TA: transcriptomic age, TAaccel: Transcriptomic Age Acceleration (transcriptomic age minus chronological age), PHQ-9: Patient Health Questionnaire 9 item depression module, PSS-10: 10 item Perceived Stress Scale, PSQI: Pittsburgh Sleep Quality Index, SMM: Skeletal Muscle Mass (lbs.), BFM: Body Fat mass (lbs.), BMI: Body Mass Index.

### Primary Analysis: Transcriptomic Age

A significant group x time difference in TA (p=.026) was observed. A significant decrease in TA was observed in the experimental group (p=.043) and a significant increase in TA was observed in the control group (p=.018) (Table 2). Changes to TAaccel were also significant between groups (p=.025), with similar magnitude and direction of change as TA (Table 2).

### Secondary Analyses: Gene Expression Analyses, Depression, Sleep, and Stress Ratings

There was a significant decrease in mean PHQ-9 (depression) and PSQI (sleep) (p=.002 and p=.042), over time for the experimental group but no significant change for control group (p=.063 and p=.158 respectively) (Table2). Lastly, there was no significant change in mean PSS-10 and SMM (p=.53 and p=.676 respectively) for the experimental group and similarly for the control group (p=.054 and p=.456). However, no changes in stress, sleep, or depression ratings displayed group x time statistical significance.

The group x time interaction gene expression analysis identified 98 genes that were differentially expressed using routinely accepted fold change cutoff values (86 up-regulated genes >1.5-fold change, and 12 down regulated genes <.5-fold change in the exercise group compared to control group). This number is insufficient for secondary enrichment analyses. Using more liberal fold change values of >1.2 and < .8 for this exploratory analysis, 2,653 DEGs were identified (1075 up-regulated genes >1.2-fold change, and 1778 down-regulated genes <.8-fold change) (Supplementary File 1). In addition, 1,365 Gene Ontology (GO) terms, 477 gene upstream regulators, 231 chemical upstream regulators and 259 diseases were found to be significantly enriched before correction for multiple comparisons (Supplementary File 2).

Pathway analysis was performed using Advaita Bio’s iPathwayGuide, which scores pathways using the Impact Analysis method (Draghici et al., 2007; Tarca et al., 2009). Impact analysis uses two types of evidence: i) the over-representation of differentially expressed (DE) genes in a pathway and ii) the perturbation of that pathway computed by propagating the measured expression changes across the pathway topology. The top five pathways identified by this analysis and their associated p-values are as follows: Human T-cell leukemia virus 1 infection (p-value= 2.033e-7, p-value (FDR)= 3.888e-5, p-value (Bonferroni)= 6.851e-5), pathways in cancer (p-value= 2.308e-7, p-value (FDR)= 3.888e-5, p-value (Bonferroni)= 7.776e-5), neurotrophin signaling pathway (p-value= 4.670e-7, p-value (FDR)= 5.246e-5, p-value (Bonferroni)= 1.574e-4), RNA degradation (p-value= 1.140e-6, p-value (FDR)= 5.939e-5, p-value (Bonferroni)= 3.842e-4), and autophagy (p-value= 1.190e-6, p-value (FDR)= 5.939e-5, p-value (Bonferroni)= 4.009e-4). A detailed description of these results, including pathway diagrams, is shown in Supplementary File 2.

## Discussion

in this randomized controlled trial examining the effects of HIIT on an RNA-based measure of biological age, participants in the HIIT group showed greater reductions in TA and TAaccel than did those in the no-exercise control group. This improvement in biological age coincided with improvements in body composition, ratings of sleep quality, and ratings of depression within the exercise group. These results suggest that exercise exerts a causal effect on age-related patterns of gene expression, and that such effects could potentially contribute to the positive health and longevity effects associated with exercise.

### Transcriptomic Age and Transcriptomic Age Acceleration

Both groups began the trial with positive transcriptomic age acceleration. In other words, mean transcriptomic age was higher than mean chronological age in both groups. In the exercise group, TA and TAaccel decreased following the HIIT protocol, while both measures increased in the control group over the same timeframe. A 3.59-year reversal of TA was observed in the exercise group, which can be interpreted as the average gene expression pattern among exercise participants changing to reflect that of a person 3.59 years younger than their mean baseline TA. The 6.87-year difference in TA, and 7.04-year difference in TAaccel between exercise and control groups was significant.

The only significant change observed in the control group was increased TA, and the authors propose two potential mechanisms for this. Control participants were asked to avoid altering their typical physical activity levels during the duration of the four-week control protocol. It is possible that once under observation, participants inadvertently lowered their activity levels. In essence, a Hawthorne effect (Merrett, 2006). Secondly, it is important to note the profound impact that loneliness and isolation can have on gene expression (Steve W. Cole, 2009; S. W. Cole et al., 2015). Many control participants expressed disappointment at not being included in the exercise group. It is at least conceivable that this adversely affected their transcriptomic age.

Of note was that the TRAP model consistently overestimated participant age in all blood samples. The authors believe this is due to differences in data type between the TRAP training dataset and our sample. The TRAP model was developed and trained using microarray data (Peters et al., 2015), while our transcript counts were derived from RNAseq data. However, since this discrepancy applies equally to all blood samples regardless of group assignment or time of collection, there is no reason to believe that this introduced any bias into the observed magnitude and direction of TA change.

### Gene Expression

The use of a gene expression-based measure of biological age has the added advantage of facilitating additional transcriptomic analyses which could shed light on the mechanisms underlying exercise’s effect on aging processes. However, in an untargeted genome-wide expression analysis, 12 HIIT sessions had only modest effects on gene expression. Although there were transcriptomic effects associated with HIIT, less than 100 genes displayed a fold change greater than 1.5 or less than .5, the values typically used to identify DEGs. This DEG count is less than the amount required for subsequent higher order bioinformatic analyses such as a functional enrichment analysis.

While these modest findings may seem surprising given the systemic physiological changes induced by exercise, it is important to remember that this trial examined the effects of a 1-month HIIT protocol on steady state expression levels. The follow up blood draw occurred approximately 48 hours after the final exercise session, meaning that whole genome expression was assessed while the participants were not experiencing the acute physiological aftereffects of exercise. Given the small dose and duration of our exercise protocol and the small sample size, this modest between group effect may not be surprising

An exploratory genome-wide discovery analysis using more liberal fold change cutoff values (greater than 1.2 or less than .8) revealed 1075 upregulated transcripts and 1778 downregulated transcripts potentially associated with HIIT (Supplementary File 1). The subsequent bioinformatic analyses associated with these DEGs were performed using Advaita Bio’s iPathwayGuide. This analysis suggests that autophagy processes, cancer pathways, neurotrophin signaling pathways, mRNA degradation processes, and other pathways were modified by HIIT (Supplementary File 2). These modifications are particularly interesting in the context of aging, especially autophagy. Various age-related signaling pathways were modified including mTOR signaling, AMPK signaling, PI3K signaling, and insulin signaling pathways. Inhibition of 3 out of 5 mTORC1 complex component genes (Raptor, Deptor, and mTOR) was noteworthy, since mTORC1 inhibition is associated with increased lifespan in every species studied so far, including humans (Papadopoli et al., 2019; Weichhart, 2018). An upstream mTORC1 activator, RHEB, was also inhibited. RHEB activates mTOR by antagonizing its endogenous inhibitor, FKBP38 (Bai et al., 2007). HIIT-induced RHEB inhibition could therefore lead to mTORC1 inhibition. Given the exploratory nature of these enrichment analyses, and the relatively liberal threshold for DEG detection however, these results should be treated as descriptive hypotheses to be tested in future research using more rigorous methods.

### Body Composition and Self-Reported Measures of Sleep Quality and Depression

Previous work suggests that the effects of exercise on biological age are mediated by changes in body composition (Kresovich et al., 2021). This seems to support our findings, as improvements in BMI, body fat mass, and visceral fat area were observed in the exercise group over time. Improvements in PHQ-9 and PSQI score were also seen in the exercise group over time.

Observed changes to body composition were consistent with previous studies’ findings, indicating that this study’s specific implementation of HIIT imparted the expected effects demonstrated in prior investigations. This serves as a positive control, or paradigm validation of the trial’s specific HIIT intervention.

### Significance

Starting and adhering to a new exercise program is difficult, a fact perhaps best illustrated by the current sedentary behavior rate in the United States. A recent Center for Disease Control and Prevention (CDC) telephone survey estimates that more than 25% of Americans participate in no physical activity outside of work (CDC, 2022) and contrary to popular opinion, this is not a uniquely American problem. A large European Union study found that 53.1% of the adult EU population participated in >4.5 hours of sedentary behavior per day (López-Valenciano et al., 2020). Inadequate physical activity is no longer just a western problem either, with the World Health Organization estimating that one third of the global population aged 15 years or older engages in insufficient physical activity, with some countries, such as Korea, engaging in >8 hours per day of sedentary behavior on average (Park, Moon, Kim, Kong, & Oh, 2020).

HIIT is a potential tool to help combat this trend given the decreased time commitment (Cobbold, 2018; Ito, 2019) and similar (or improved) health benefits to those bestowed by other forms of exercise (Hannan et al., 2018; Scott et al., 2019), but with increased adherence and compliance rates (Ito, 2019).

Despite the modest gene expression findings generally, the pre-specified hypothesis regarding HIIT-induced transcriptomic age reversal was proven out by the analysis. Considering that each exercise participant completed a combined 276 minutes of exercise over 1 month, only 2 hours of which was high intensity exercise, the effect of HIIT on biological age appears promising.

This study further supports the notion that adding even a small amount of exercise can be beneficial, given that just 12 HIIT sessions were shown to significantly improve TA and TAaccel. To the authors’ knowledge, this is the first trial to demonstrate the effects of a specific exercise protocol on a next generation measure of biological age. The results suggest that exercise exerts a causal effect on age-related patterns of gene expression, and that such effects could potentially contribute to the positive health and longevity effects associated with exercise.

## Conclusion

A low dose of HIIT is sufficient to reduce transcriptomic age in sedentary middle-aged males and females. Other changes to gene expression were relatively modest in comparison to the transcriptomic age reversal effect size. These findings, along with modification to autophagic pathways, may indicate a particular HIIT specificity for age-related biological pathway modulation. The key observations presented here, namely reduced transcriptomic age, indicate that exercise may potentially improve health and longevity by altering age-related transcriptional processes.

## Data Availability

The data that support the findings of this study are available from the corresponding author, [T.L.], upon reasonable request.

## Acknowledgements

We are grateful for the work carried out by our laboratory team: Chih-Chieh Chia, Fulden Cakir, Simran Jaisinghani, Kezia Marceline, Owee Mulay, and Maxine Shih. We also thank this paper’s reviewers, who improved the manuscript in meaningful ways.

## Permission Statement

### Authors’ Contributions

T.L. conceived and designed the study, performed transcriptomic age and associated enrichment analyses, G.B. coordinated the project and together with T.L. and E.L. designed the study. S.C. performed all RNA extraction, quality control, and associated transcriptomic analyses. L.G. performed associated statistical analyses. All authors contributed to and approved of the manuscript.

