## Supplementary File 2 for "High-Intensity Interval Training Reduces Transcriptomic Age: A Randomized Controlled Trial"

iPathwayGuide

| | |
|---|---|
| **Title:** | The Effects of High Intensity Interval Training on Gene Expression |
| **Description:** | Effects of a 3 times per week, 4-week, 10X1 HIIT protocol on gene expression. Functional Enrichment analysis cutoff threshold >1.2 or <.8 |
| **Organism:** | Homo sapiens (9606) |
| **Contrast** | Condition vs. Control - mRNA (RNA-seq) |
| **Creation time:** | 10-23-2022 06:42 PM |

### 1. Introduction

In this experiment, **2,653** differentially expressed (DE) genes were identified out of a total of **54,683** genes in Advaita Knowledge Base (AKB). These data were analyzed in the context of pathways obtained from the Kyoto Encyclopedia of Genes and Genomes (KEGG) database (Release 100.0+/11-12, Nov 21) (Kanehisa et al., 2000; Kanehisa et al., 2002), gene ontologies from the Gene Ontology Consortium database (2021-Nov4) (Ashburner et al., 2000; Gene Ontology Consortium, 2001), miRNAs from the miRBase (MIRBASE Version:Version22.1,10/18) and TARGETSCAN (Targetscan version: Mouse:8.0, Human:8.0) databases (Agarwal et al., 2015; Nam et al., 2014; Griffiths-Jones et al., 2008; Kozomara and Griffiths-Jones, 2014; Friedman et al., 2009; Grimson et al., 2007), network of regulatory relations from BioGRID: Biological General Repository for Interaction Datasets v4.4.203. Oct. 25th, 2021 (Szklarczyk et al., 2017), chemicals/drugs/toxicants from the Comparative Toxicogenomics Database Nov 2021 (Davis et al., 2019), and diseases from the KEGG database (Release 100.0+/11-12, Nov 21) (Kanehisa et al., 2000; Kanehisa et al., 2002). In summary, **229** pathways were found to be significantly impacted. In addition, **1,365** Gene Ontology (GO) terms, **0** miRNAs , **477** gene upstream regulators, **231** chemical upstream regulators and **259** diseases were found to be significantly enriched before the correction for multiple comparisons.

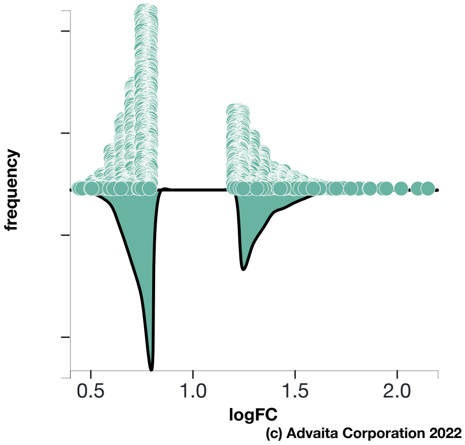


a)

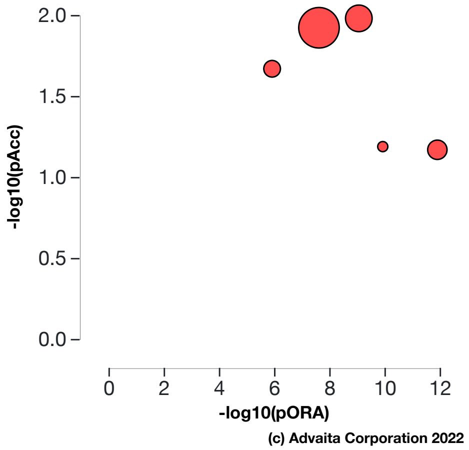


b)

**Fig. 1.1: a) Violin plot:** *All **2653** significantly differentially expressed (DE) genes are represented in terms of their measured expression change (x-axis) and frequency of genes measured at a given expression change (y-axis)* **b) Pathways perturbation vs over-representation:** *The top 5 pathways are plotted in terms of the two types of evidence computed by iPathwayGuide: over-representation on the x-axis (pORA) and the total pathway accumulation on the y-axis (pAcc). Each pathway is represented by a single dot, with significant pathways shown in red, non-significant in black, and the size of each dot is proportional to the size of the pathway it represents. Both p-values are shown in terms of their negative log (base 10) values.*

### 2. Pathway Analysis

#### 2.1. Methods

iPathwayGuide scores pathways using the Impact Analysis method (Draghici et al., 2007; Tarca et al., 2009, Khatri et al., 2007). Impact analysis uses two types of evidence: i) the over-representation of differentially expressed (DE) genes in a given pathway and ii) the perturbation of that pathway computed by propagating the measured expression changes across the pathway topology. These aspects are captured by two independent probability values, pORA and pAcc, that are then combined in a unique pathway-specific p-value. The underlying pathway topologies, comprised of genes and their directional interactions, are obtained from the KEGG database (Kanehisa et al., 2000; Kanehisa et al., 2010; Kanehisa et al., 2012; Kanehisa et al., 2014).

The first probability, pORA, expresses the probability of observing the number of DE genes in a given pathway that is greater than or equal to the number observed, by random chance (Draghici et al., 2003; Draghici 2011). Let us consider there are *N* genes measured in the experiment, with *M* of these on the given pathway. Based on the user-defined a priori selection of DE genes, *K* out of *M* genes were found to be differentially expressed. The probability of observing exactly *x* differentially expressed genes on the given pathway is computed based on the hypergeometric distribution:

$$(1) \qquad P(X=x|N,M,K) = \frac{\binom{M}{x}\binom{N-M}{K-x}}{\binom{N}{K}}$$

Because the hypergeometric distribution is discrete, the probability of observing fewer than *x* genes on the given pathway just by chance can be calculated by summing the probabilities of randomly observing *0*, *1*, *2*, ..., up to *x-1* DE genes on the pathway:

$$(2) \qquad p_u(x-1) = P(X=1)+P(X=2)+\ldots+P(X=x-1) = \sum_{i=0}^{x-1} \frac{\binom{M}{i}\binom{N-M}{K-i}}{\binom{N}{K}}$$

iPathwayGuide calculates the probability of randomly observing a number of DE genes on the given pathway that is greater than or equal to the number of DE genes obtained from data, by computing the over-representation p-value: $pORA = p_o(x) = 1 - p_u(x-1)$:

$$(3) \qquad p_o(x) = 1 - \sum_{i=0}^{x-1} \frac{\binom{M}{i}\binom{N-M}{K-i}}{\binom{N}{K}}$$

The second probability, pAcc, is calculated based on the amount of total accumulation measured in each pathway. A perturbation factor is computed for each gene on the pathway using:

$$(4) \qquad PF(g) = \alpha(g) \cdot \triangle E(g) + \sum_{u \in US_g} \beta_{ug} \frac{PF(u)}{N_{ds}(u)}$$

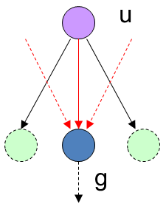


In Equation 4, *PF(g)* is the perturbation factor for gene *g*, the term *ΔE(g)* represents the signed normalized measured expression change of gene *g*, and *α(g)* is a priori weight based on the type of the gene. The last term is the sum of the perturbation factors of all genes *u*, directly upstream of the target gene *g*, normalized by the number of downstream genes of each such gene $N_{ds}(u)$. The value of $\beta_{ug}$ quantifies the strength of the interaction between genes *g* and *u*. The sign of *β* represents the type of interaction: positive for activation-like signals, and negative for inhibition-like signals. Subsequently, iPathwayGuide calculates the accumulation at the level of each gene, *Acc(g)*, as the difference between the perturbation factor *PF(g)* and the observed log fold-change:

$$(5) \qquad Acc(g_i) = PF(g_i) - \triangle E(g_i)$$

All perturbation accumulations are computed at the same time by solving the system of linear equations resulting from combining Equation 4 for all genes on a given pathway. Once all gene perturbation accumulations are computed, iPathwayGuide computes the total accumulation of the pathway as the sum of all absolute accumulations of the genes in a given pathway. The significance of obtaining a total accumulation (pAcc) at least as large as observed, just by chance, is assessed through bootstrap analysis.

The two types of evidence, pORA and pAcc, are combined into an overall pathway score by calculating a p-value using Fisher's method. This p-value is then corrected for multiple comparisons using false dicovery rate (FDR) and Bonferroni corrections. Bonferroni is simpler and more conservative of the two (Bonferroni, 1935; Bonferroni, 1936). It reduces the false discovery rate by imposing a stringent threshold on each comparison adjusted for the total number of comparisons. The FDR correction has more power, but only controls the family-wise false positives rate (Benjamini and Hochberg, 1995; Benjamini and Yekutieli, 2001).

### 2.2. Results

**Table 2.2.1: Top pathways and their associated p-values**

| Pathway name | Pathway Id | p-value | p-value (FDR) | p-value (Bonferroni) |
|---|---|---|---|---|
| Human T-cell leukemia virus 1 infection | 05166 | **2.033e-7** | **3.888e-5** | **6.851e-5** |
| Pathways in cancer | 05200 | **2.308e-7** | **3.888e-5** | **7.776e-5** |
| Neurotrophin signaling pathway | 04722 | **4.670e-7** | **5.246e-5** | **1.574e-4** |
| RNA degradation | 03018 | **1.140e-6** | **5.939e-5** | **3.842e-4** |
| Autophagy - animal | 04140 | **1.190e-6** | **5.939e-5** | **4.009e-4** |

* the p-value corresponding to the pathway was computed using only over-representation analysis.

#### Human T-cell leukemia virus 1 infection (KEGG: 05166)

Human T-cell leukemia virus type 1 (HTLV-1) is a pathogenic retrovirus that is associated with adult T-cell leukemia/lymphoma (ATL). It is also strongly implicated in non-neoplastic chronic inflammatory diseases such as HTLV-1-associated myelopathy/tropical spastic paraparesis (HAM/TSP). Expression of Tax, a viral regulatory protein is critical to the pathogenesis. Tax is a transcriptional co-factor that interfere several signaling pathways related to anti-apoptosis or cell proliferation. The modulation of the signaling by Tax involve its binding to transcription factors like CREB/ATF, NF-kappa B, and SRF.

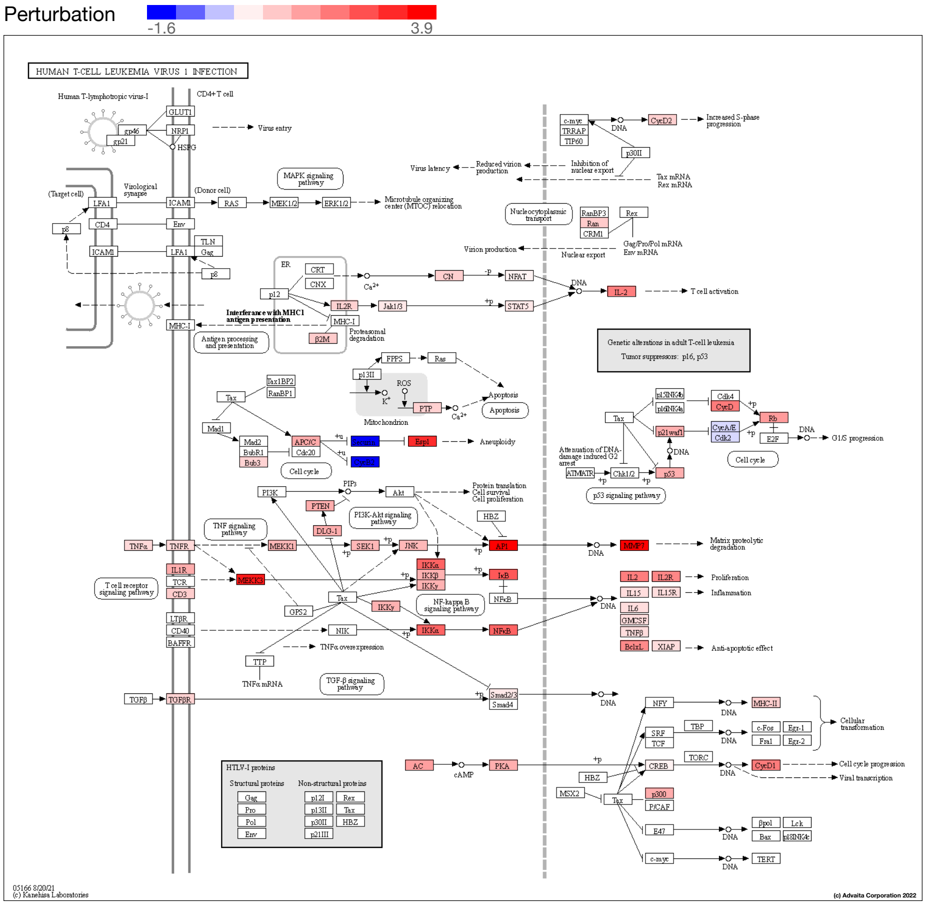


**Fig. 2.2.1: Human T-cell leukemia virus 1 infection (KEGG: 05166):** *The pathway diagram is overlayed with the computed perturbation of each gene. The perturbation accounts both for the gene's measured fold change and for the accumulated perturbation propagated from any upstream genes (accumulation). The highest negative perturbation is shown in dark blue, while the highest positive perturbation in dark red. The legend describes the values on the gradient. Note: For legibility, one gene may be represented in multiple places in the diagram and one box may represent multiple genes in the same gene family. A gene is highlighted in all locations it occurs in the diagram. For each gene family, the color corresponding to the gene with the highest absolute perturbation is displayed.*

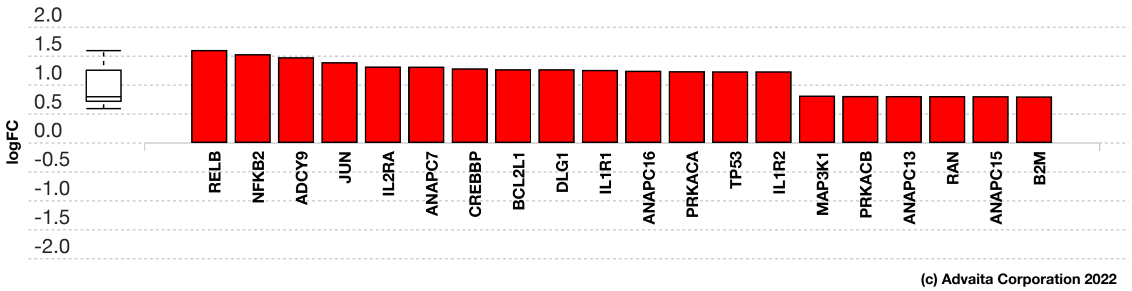


**Fig. 2.2.2: Gene measured expression bar plot:** *All the differentially expressed genes in Human T-cell leukemia virus 1 infection (KEGG: 05166) are ranked based on their absolute value of log fold change. The plot is limited to the top 20 genes out of a total of 35 differentially expressed genes. Upregulated genes are shown in red, downregulated genes are shown in blue. The box and whisker plot on the left summarizes the distribution of all the differentially expressed genes in this pathway. The box represents the 1st quartile, the median and the 3rd quartile, while the outliers are represented by circles.*

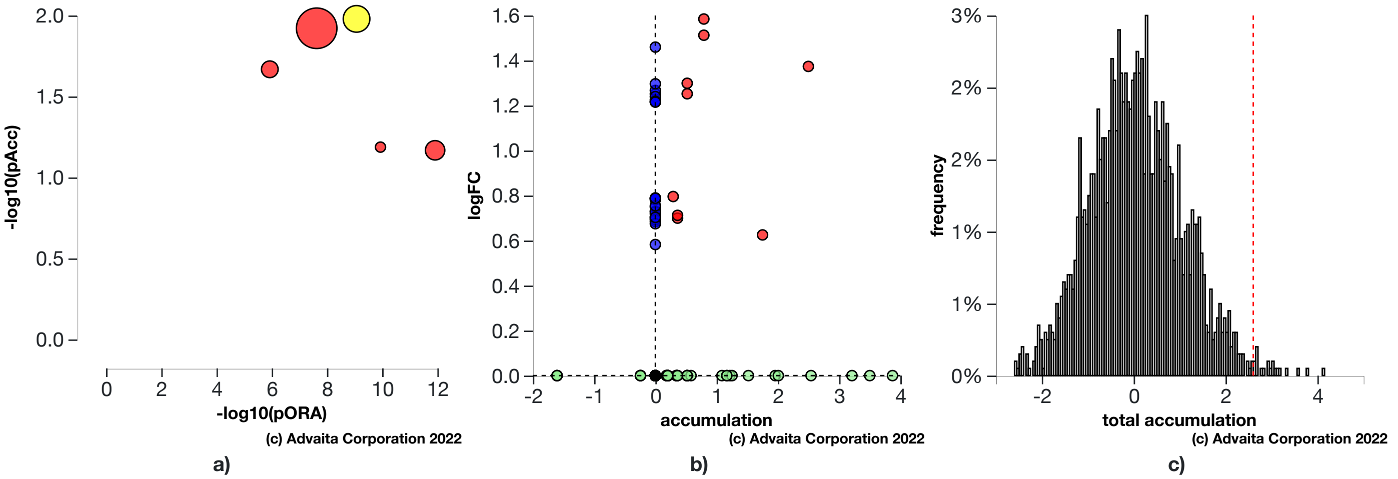


**Fig. 2.2.3: a) Perturbation vs over-representation:** *Human T-cell leukemia virus 1 infection (KEGG: 05166) (yellow) is shown, using negative log of the accumulation and over-representation p-values, along with the other most significant pathways. Pathways in red are significant based on the combined uncorrected p-values, whereas the ones in black are non-significant (where applicable).* **b) Gene measured expression vs accumulation:** *All the genes from this pathway are represented in terms of their measured fold change (y-axis) and accumulation (x-axis). Accumulation is the perturbation received by the gene from any upstream genes. Genes displayed in red had both accumulation and measured fold change.Genes in blue had only measured fold change.Genes in green had only accumulation.The remaining genes that were not measured and had no accumulation are shown in black.* **c) Bootstrap diagram:** *The perturbation p-value is computed using bootstrap analysis. Bootstrapping assesses the probability of observing a sum of all absolute gene accumulation total accumulation at least as extreme as the computed one just by chance. A null distribution (gray bars) is computed through an iterative process that is repeated 2000 times. At each iteration, a number of genes equal to the number of differentially expressed genes in this pathway is randomly assigned anywhere in the pathway and the total accumulation is recomputed. The red line indicates the observed total accumulation of genes in the given pathway in relation to the distribution of expected values. The perturbation p-value is more significant the further away from the mean it is.*

#### Pathways in cancer (KEGG: 05200)

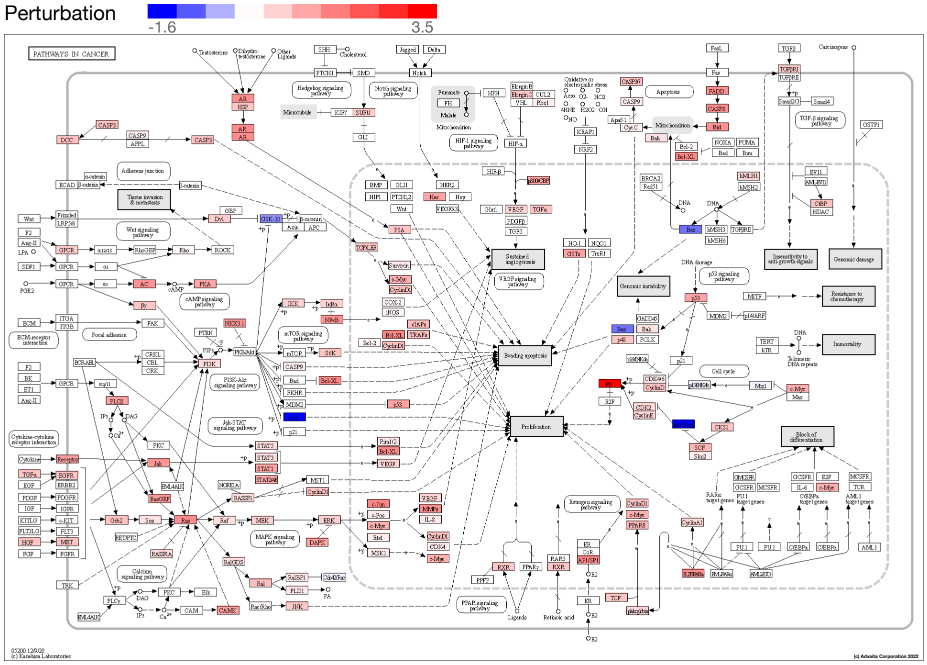


**Fig. 2.2.4: Pathways in cancer (KEGG: 05200):** *The pathway diagram is overlayed with the computed perturbation of each gene. The perturbation accounts both for the gene's measured fold change and for the accumulated perturbation propagated from any upstream genes (accumulation). The highest negative perturbation is shown in dark blue, while the highest positive perturbation in dark red. The legend describes the values on the gradient. Note: For legibility, one gene may be represented in multiple places in the diagram and one box may represent multiple genes in the same gene family. A gene is highlighted in all locations it occurs in the diagram. For each gene family, the color corresponding to the gene with the highest absolute perturbation is displayed.*

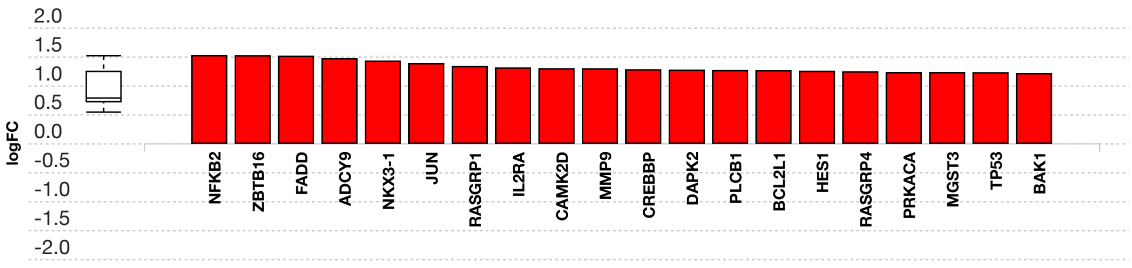


**(c) Advaita Corporation 2022**

**Fig. 2.2.5: Gene measured expression bar plot:** *All the differentially expressed genes in Pathways in cancer (KEGG: 05200) are ranked based on their absolute value of log fold change. The plot is limited to the top 20 genes out of a total of 57 differentially expressed genes. Upregulated genes are shown in red, downregulated genes are shown in blue. The box and whisker plot on the left summarizes the distribution of all the differentially expressed genes in this pathway. The box represents the 1st quartile, the median and the 3rd quartile, while the outliers are represented by circles.*

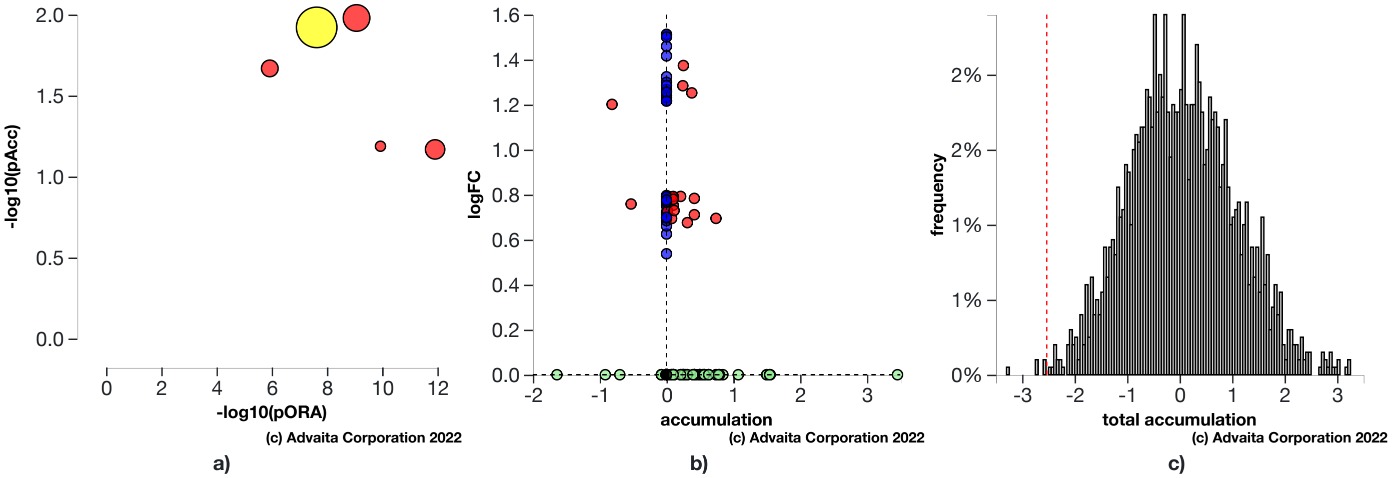


**Fig. 2.2.6: a) Perturbation vs over-representation:** *Pathways in cancer (KEGG: 05200) (yellow) is shown, using negative log of the accumulation and over-representation p-values, along with the other most significant pathways. Pathways in red are significant based on the combined uncorrected p-values, whereas the ones in black are non-significant (where applicable).* **b) Gene measured expression vs accumulation:** *All the genes from this pathway are represented in terms of their measured fold change (y-axis) and accumulation (x-axis). Accumulation is the perturbation received by the gene from any upstream genes. Genes displayed in red had both accumulation and measured fold change.Genes in blue had only measured fold change.Genes in green had only accumulation.The remaining genes that were not measured and had no accumulation are shown in black.* **c) Bootstrap diagram:** *The perturbation p-value is computed using bootstrap analysis. Bootstrapping assesses the probability of observing a sum of all absolute gene accumulation total accumulation at least as extreme as the computed one just by chance. A null distribution (gray bars) is computed through an iterative process that is repeated 2000 times. At each iteration, a number of genes equal to the number of differentially expressed genes in this pathway is randomly assigned anywhere in the pathway and the total accumulation is recomputed. The red line indicates the observed total accumulation of genes in the given pathway in relation to the distribution of expected values. The perturbation p-value is more significant the further away from the mean it is.*

#### Neurotrophin signaling pathway (KEGG: 04722)

Neurotrophins are a family of trophic factors involved in differentiation and survival of neural cells. The neurotrophin family consists of nerve growth factor (NGF), brain derived neurotrophic factor (BDNF), neurotrophin 3 (NT-3), and neurotrophin 4 (NT-4). Neurotrophins exert their functions through engagement of Trk tyrosine kinase receptors or p75 neurotrophin receptor (p75NTR). Neurotrophin/Trk signaling is regulated by connecting a variety of intracellular signaling cascades, which include MAPK pathway, PI-3 kinase pathway, and PLC pathway, transmitting positive signals like enhanced survival and growth. On the other hand, p75NTR transmits both positive and nagative signals. These signals play an important role for neural development and additional higher-order activities such as learning and memory.

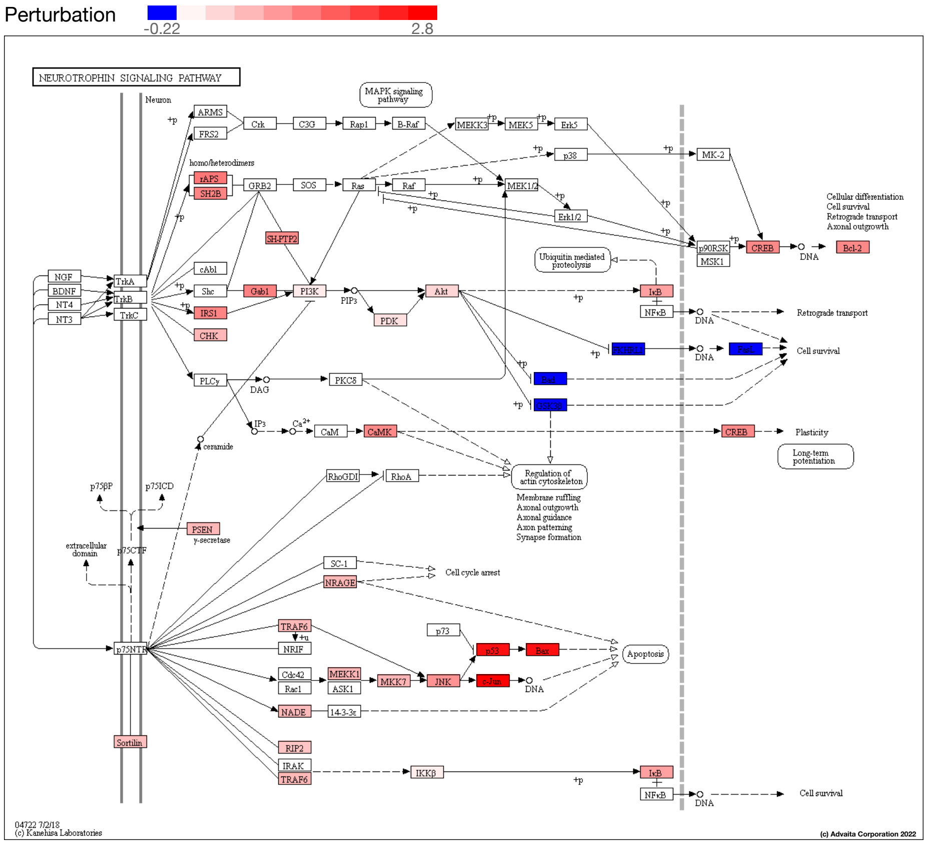


**Fig. 2.2.7: Neurotrophin signaling pathway (KEGG: 04722):** *The pathway diagram is overlayed with the computed perturbation of each gene. The perturbation accounts both for the gene's measured fold change and for the accumulated perturbation propagated from any upstream genes (accumulation). The highest negative perturbation is shown in dark blue, while the highest positive perturbation in dark red. The legend describes the values on the gradient. Note: For legibility, one gene may be represented in multiple places in the diagram and one box may represent multiple genes in the same gene family. A gene is highlighted in all locations it occurs in the diagram. For each gene family, the color corresponding to the gene with the highest absolute perturbation is displayed.*

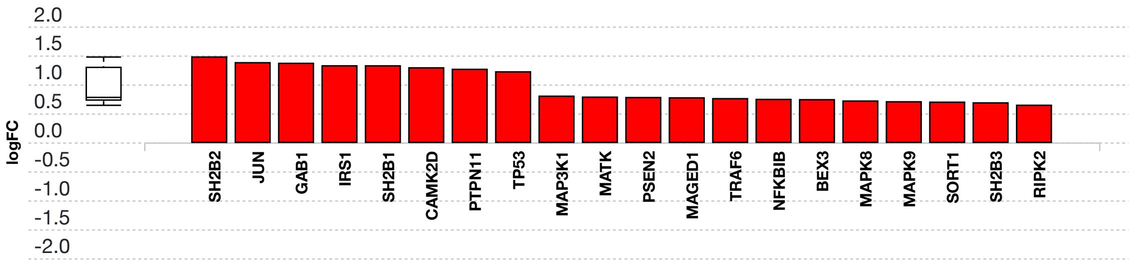


**Fig. 2.2.8: Gene measured expression bar plot:** *All the differentially expressed genes in Neurotrophin signaling pathway (KEGG: 04722) are ranked based on their absolute value of log fold change. Upregulated genes are shown in red, downregulated genes are shown in blue. The box and whisker plot on the left summarizes the distribution of all the differentially expressed genes in this pathway. The box represents the 1st quartile, the median and the 3rd quartile, while the outliers are represented by circles.*

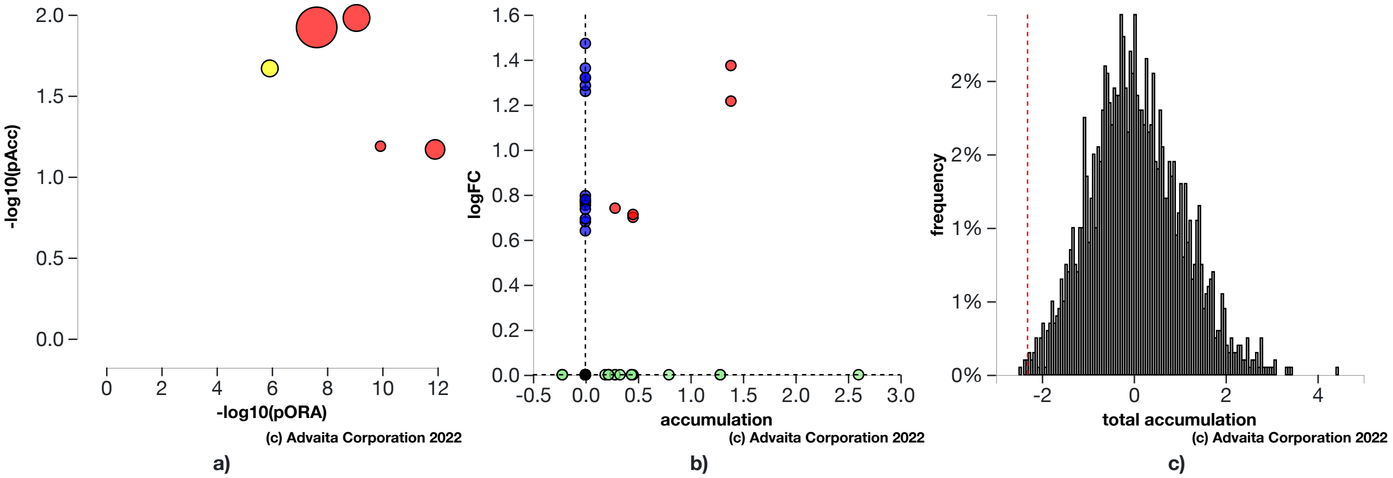


**Fig. 2.2.9: a) Perturbation vs over-representation:** *Neurotrophin signaling pathway (KEGG: 04722) (yellow) is shown, using negative log of the accumulation and over-representation p-values, along with the other most significant pathways. Pathways in red are significant based on the combined uncorrected p-values, whereas the ones in black are non-significant (where applicable).* **b) Gene measured expression vs accumulation:** *All the genes from this pathway are represented in terms of their measured fold change (y-axis) and accumulation (x-axis). Accumulation is the perturbation received by the gene from any upstream genes. Genes displayed in red had both accumulation and measured fold change.Genes in blue had only measured fold change.Genes in green had only accumulation.The remaining genes that were not measured and had no accumulation are shown in black.* **c) Bootstrap diagram:** *The perturbation p-value is computed using bootstrap analysis. Bootstrapping assesses the probability of observing a sum of all absolute gene accumulation total accumulation at least as extreme as the computed one just by chance. A null distribution (gray bars) is computed through an iterative process that is repeated 2000 times. At each iteration, a number of genes equal to the number of differentially expressed genes in this pathway is randomly assigned anywhere in the pathway and the total accumulation is recomputed. The red line indicates the observed total accumulation of genes in the given pathway in relation to the distribution of expected values. The perturbation p-value is more significant the further away from the mean it is.*

#### RNA degradation (KEGG: 03018)

The correct processing, quality control and turnover of cellular RNA molecules are critical to many aspects in the expression of genetic information. In eukaryotes, two major pathways of mRNA decay exist and both pathways are initiated by poly(A) shortening of the mRNA. In the 5' to 3' pathway, this is followed by decapping which then permits the 5' to 3' exonucleolytic degradation of transcripts. In the 3' to 5' pathway, the exosome, a large multisubunit complex, plays a key role. The exosome exists in archaeal cells, too. In bacteria, endoribonuclease E, a key enzyme involved in RNA decay and processing, organizes a protein complex called degradosome. RNase E or R interacts with the phosphate-dependent exoribonuclease polynucleotide phosphorylase, DEAD-box helicases, and additional factors in the RNA-degrading complex.

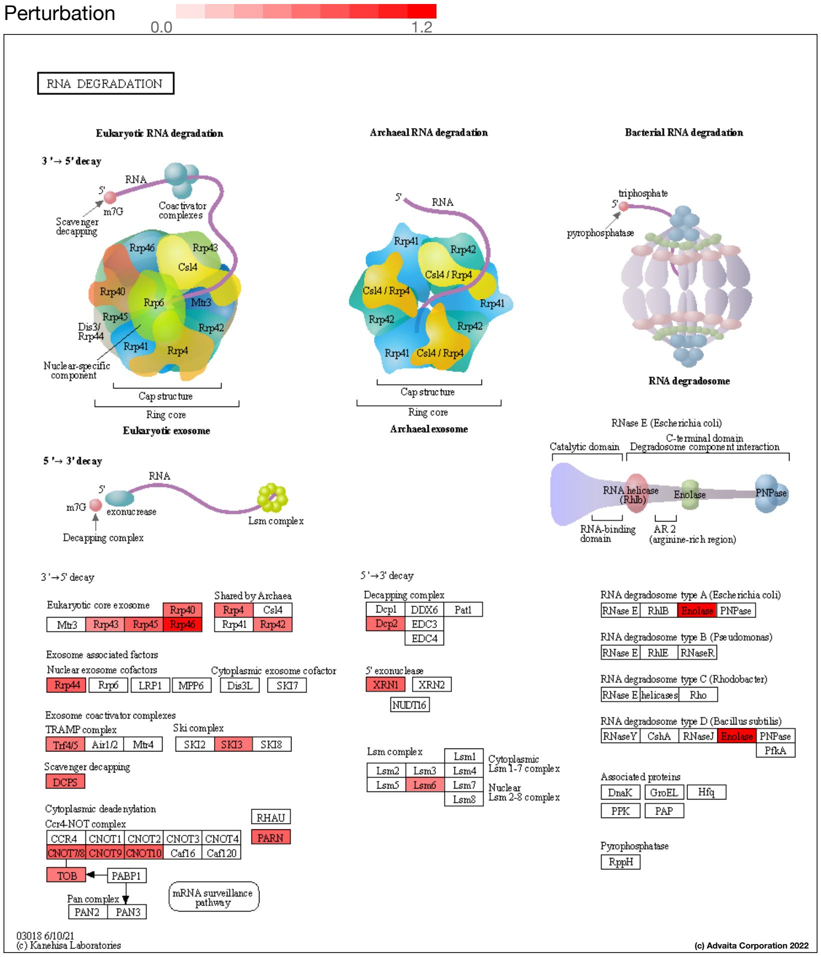


**Fig. 2.2.10: RNA degradation (KEGG: 03018):** *The pathway diagram is overlayed with the computed perturbation of each gene. The perturbation accounts both for the gene's measured fold change and for the accumulated perturbation propagated from any upstream genes (accumulation). The highest negative perturbation is shown in dark blue, while the highest positive perturbation in dark red. The legend describes the values on the gradient. Note: For legibility, one gene may be represented in multiple places in the diagram and one box may represent multiple genes in the same gene family. A gene is highlighted in all locations it occurs in the diagram. For each gene family, the color corresponding to the gene with the highest absolute perturbation is displayed.*

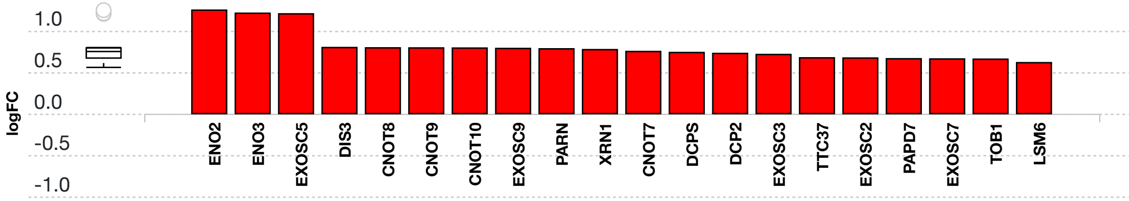


(c) Advaita Corporation 2022

**Fig. 2.2.11: Gene measured expression bar plot:** *All the differentially expressed genes in RNA degradation (KEGG: 03018) are ranked based on their absolute value of log fold change. The plot is limited to the top 20 genes out of a total of 21 differentially expressed genes. Upregulated genes are shown in red, downregulated genes are shown in blue. The box and whisker plot on the left summarizes the distribution of all the differentially expressed genes in this pathway. The box represents the 1st quartile, the median and the 3rd quartile, while the outliers are represented by circles.*

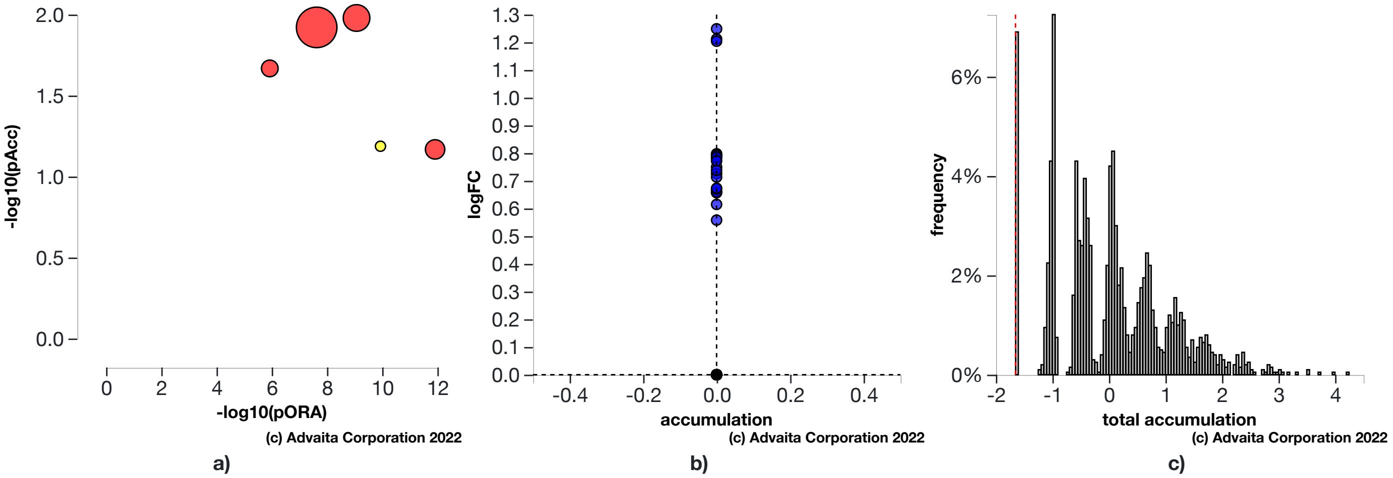


**Fig. 2.2.12: a) Perturbation vs over-representation:** *RNA degradation (KEGG: 03018) (yellow) is shown, using negative log of the accumulation and over-representation p-values, along with the other most significant pathways. Pathways in red are significant based on the combined uncorrected p-values, whereas the ones in black are non-significant (where applicable).* **b) Gene measured expression vs accumulation:** *All the genes from this pathway are represented in terms of their measured fold change (y-axis) and accumulation (x-axis). Accumulation is the perturbation received by the gene from any upstream genes. Genes in blue had only measured fold change.The remaining genes that were not measured and had no accumulation are shown in black.* **c) Bootstrap diagram:** *The perturbation p-value is computed using bootstrap analysis. Bootstrapping assesses the probability of observing a sum of all absolute gene accumulation total accumulation at least as extreme as the computed one just by chance. A null distribution (gray bars) is computed through an iterative process that is repeated 2000 times. At each iteration, a number of genes equal to the number of differentially expressed genes in this pathway is randomly assigned anywhere in the pathway and the total accumulation is recomputed. The red line indicates the observed total accumulation of genes in the given pathway in relation to the distribution of expected values. The perturbation p-value is more significant the further away from the mean it is.*

#### Autophagy - animal (KEGG: 04140)

Autophagy (or macroautophagy) is a cellular catabolic pathway involving in protein degradation, organelle turnover, and non-selective breakdown of cytoplasmic components, which is evolutionarily conserved among eukaryotes and exquisitely regulated. This progress initiates with production of the autophagosome, a double-membrane intracellular structure of reticular origin that engulfs cytoplasmic contents and ultimately fuses with lysosomes for cargo degradation. Autophagy is regulated in response to extra- or intracellular stress and signals such as starvation, growth factor deprivation and ER stress. Constitutive level of autophagy plays an important role in cellular homeostasis and maintains quality control of essential cellular components.

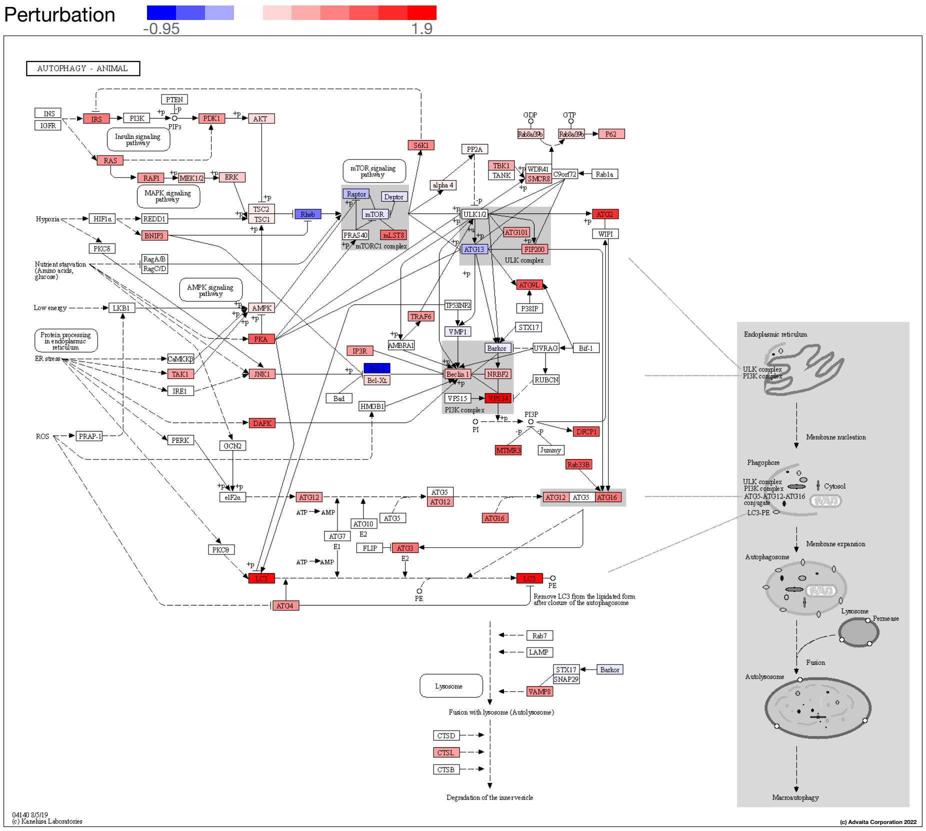


**Fig. 2.2.13: Autophagy - animal (KEGG: 04140):** *The pathway diagram is overlayed with the computed perturbation of each gene. The perturbation accounts both for the gene's measured fold change and for the accumulated perturbation propagated from any upstream genes (accumulation). The highest negative perturbation is shown in dark blue, while the highest positive perturbation in dark red. The legend describes the values on the gradient. Note: For legibility, one gene may be represented in multiple places in the diagram and one box may represent multiple genes in the same gene family. A gene is highlighted in all locations it occurs in the diagram. For each gene family, the color corresponding to the gene with the highest absolute perturbation is displayed.*

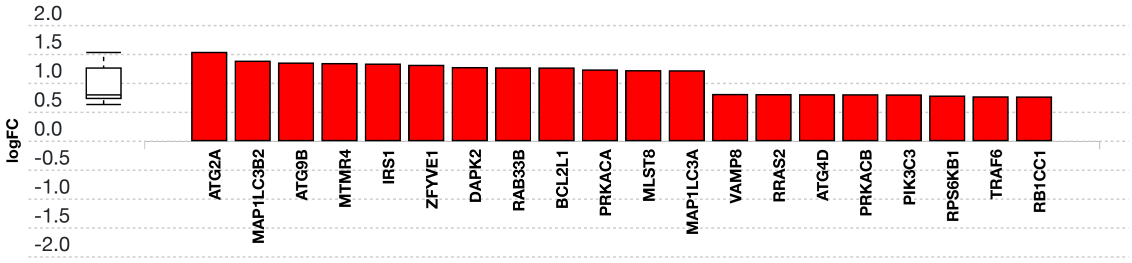


**(c) Advaita Corporation 2022**

**Fig. 2.2.14: Gene measured expression bar plot:** *All the differentially expressed genes in Autophagy - animal (KEGG: 04140) are ranked based on their absolute value of log fold change. The plot is limited to the top 20 genes out of a total of 31 differentially expressed genes. Upregulated genes are shown in red, downregulated genes are shown in blue. The box and whisker plot on the left summarizes the distribution of all the differentially expressed genes in this pathway. The box represents the 1st quartile, the median and the 3rd quartile, while the outliers are represented by circles.*

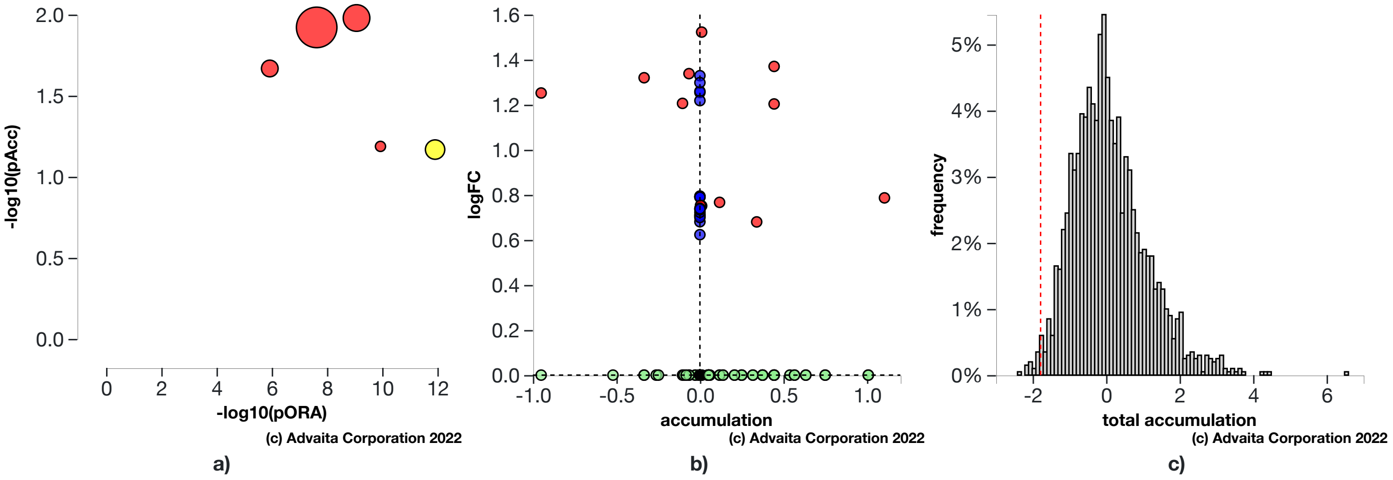


**Fig. 2.2.15: a) Perturbation vs over-representation:** *Autophagy - animal (KEGG: 04140) (yellow) is shown, using negative log of the accumulation and over-representation p-values, along with the other most significant pathways. Pathways in red are significant based on the combined uncorrected p-values, whereas the ones in black are non-significant (where applicable).* **b) Gene measured expression vs accumulation:** *All the genes from this pathway are represented in terms of their measured fold change (y-axis) and accumulation (x-axis). Accumulation is the perturbation received by the gene from any upstream genes. Genes displayed in red had both accumulation and measured fold change.Genes in blue had only measured fold change.Genes in green had only accumulation.The remaining genes that were not measured and had no accumulation are shown in black.* **c) Bootstrap diagram:** *The perturbation p-value is computed using bootstrap analysis. Bootstrapping assesses the probability of observing a sum of all absolute gene accumulation total accumulation at least as extreme as the computed one just by chance. A null distribution (gray bars) is computed through an iterative process that is repeated 2000 times. At each iteration, a number of genes equal to the number of differentially expressed genes in this pathway is randomly assigned anywhere in the pathway and the total accumulation is recomputed. The red line indicates the observed total accumulation of genes in the given pathway in relation to the distribution of expected values. The perturbation p-value is more significant the further away from the mean it is.*

### 3. Gene Ontology Analysis

#### 3.1. Methods

For each Gene Ontology (GO) term (Ashburner et al., 2002; Gene Ontology Consortium, 2004), the number of differentially expressed (DE) genes annotated to the term is compared to the number of DE genes expected just by chance. iPathwayGuide uses an over-representation approach to compute the statistical significance of observing at least the given number of DE genes. The p-value is computed using the hypergeometric distribution as described for pORA in the Pathway Analysis section. This p-value is corrected for multiple comparisons using FDR and Bonferroni.

The classical enrichment method used above considers all GO terms to be independent. By definition, all genes annotated to a GO term are also annotated to its ancestors. Because of this, the enrichment approach counts each gene multiple times by propagating it through the GO hierarchy from the most specific term the gene is associated with, all the way to the root of the ontology. This introduces redundancy in the analysis and reports many general and non-informative terms as significant. To overcome this limitation, iPathwayGuide allows users to use two more sophisticated pruning methods: *high-specificity pruning* and *smallest common denominator pruning*. The **high-specificity** pruning method *identifies the most specific GO terms* that are significantly associated with the set of DE genes. Let us consider, BP1 = “induction of apoptosis by intracellular signals” and BP2 = “induction of apoptosis by extracellular signals,” which are two of the children of BP3 = “induction of apoptosis.” If enough DE genes are associated with BP1 and BP2, the high-specificity pruning will report them as significant. The **smallest common denominator** pruning method *identifies the GO terms that best encapsulate the set of DE genes*, at times consolidating significance of two or more specific terms into their common parent. In the example above, this pruning method might report BP3 as significant because it is the most specific biological term that would include all DE genes that make both BP1 and BP2 significant.

### 3.2. Biological Processes results

**Table 3.2.1: Top identified biological processes. Only the top scoring biological process for each pruning type is described below the table.**

| Pruning Type: None | | | | Pruning Type: High-specificity | | Pruning Type: Smallest Common Denominator | |
|---|---|---|---|---|---|---|---|
| **GO Term** | **p-value** | **p-value (FDR)** | **p-value (Bonferroni)** | **GO Term** | **p-value** | **GO Term** | **p-value** |
| cellular metabolic process | **4.100e-19** | **3.769e-15** | **3.769e-15** | exonucleolytic catabolism of deadenylated mRNA | **5.883e-5** | exonucleolytic catabolism of deadenylated mRNA | **5.883e-5** |
| organonitrogen compound metabolic process | **3.600e-15** | **1.655e-11** | **3.309e-11** | nuclear-transcribed mRNA catabolic process, exonucleolytic, 3'-5' | **0.029** | tRNA processing | **0.004** |
| nitrogen compound metabolic process | **1.400e-14** | **4.290e-11** | **1.287e-10** | U4 snRNA 3'-end processing | 0.248 | RNA methylation | **0.006** |
| cellular macromolecule metabolic process | **2.100e-14** | **4.826e-11** | **1.930e-10** | nuclear polyadenylation-dependent rRNA catabolic process | 0.294 | nuclear-transcribed mRNA catabolic process, exonucleolytic, 3'-5' | **0.015** |
| macromolecule modification | **2.300e-13** | **3.677e-10** | **2.114e-9** | nuclear polyadenylation-dependent tRNA catabolic process | 0.294 | ribosome biogenesis | **0.034** |

#### cellular metabolic process (GO:0044237)

The chemical reactions and pathways by which individual cells transform chemical substances. In this experiment, the algorithm identified **1,540** differentially expressed gene(s) out of ALL **10,861** gene(s).

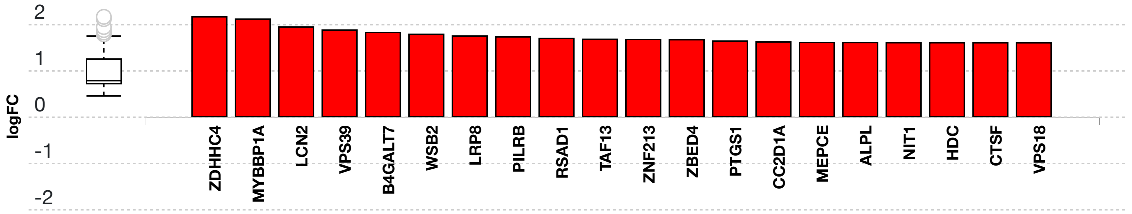


(c) Advaita Corporation 2022

**Fig. 3.2.1: Gene measured expression bar plot:** *All the differentially expressed genes that are annotated to cellular metabolic process are ranked based on their absolute value of log fold change. The plot is limited to the top 20 genes out of a total of 1540 differentially expressed genes. Upregulated genes are shown in red, downregulated genes are shown in blue. The box and whisker plot on the left summarizes the distribution of all the differentially expressed genes that are annotated to this GO term. The box represents the 1st quartile, the median and the 3rd quartile, while the outliers are represented by circles.*

#### exonucleolytic catabolism of deadenylated mRNA (GO:0043928)

The chemical reactions and pathways resulting in the breakdown of the transcript body of a nuclear-transcribed mRNA that occurs when the ends are not protected by the 3'-poly(A) tail. In this experiment, the algorithm identified **11** differentially expressed gene(s) out of ALL **13** gene(s).

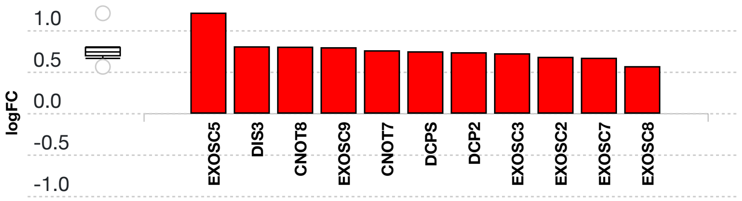


155 (c) Advaita Corporation 2022

**Fig. 3.2.2: Gene measured expression bar plot:** *All the differentially expressed genes that are annotated to exonucleolytic catabolism of deadenylated mRNA are ranked based on their absolute value of log fold change. Upregulated genes are shown in red, downregulated genes are shown in blue. The box and whisker plot on the left summarizes the distribution of all the differentially expressed genes that are annotated to this GO term. The box represents the 1st quartile, the median and the 3rd quartile, while the outliers are represented by circles.*

### 3.3. Molecular Functions results

**Table 3.3.1: Top identified molecular functions. Only the top scoring molecular function for each pruning type is described below the table.**

| Pruning Type: None | | | | Pruning Type: High-specificity | | Pruning Type: Smallest Common Denominator | |
|---|---|---|---|---|---|---|---|
| GO Term | p-value | p-value (FDR) | p-value (Bonferroni) | GO Term | p-value | GO Term | p-value |
| catalytic activity | 1.500e-18 | 2.183e-15 | 3.447e-15 | protein binding | 1.907e-9 | protein binding | 1.769e-12 |
| protein binding | 1.900e-18 | 2.183e-15 | 4.366e-15 | guanyl-nucleotide exchange factor activity | 0.010 | 3'-5' exonuclease activity | 0.003 |
| catalytic activity, acting on a nucleic acid | 3.100e-10 | 2.375e-7 | 7.124e-7 | RNA binding | 0.041 | guanyl-nucleotide exchange factor activity | 0.007 |
| transferase activity | 2.700e-9 | 1.551e-6 | 6.205e-6 | 3'-5'-exoribonuclease activity | 0.155 | exoribonuclease activity | 0.007 |
| catalytic activity, acting on RNA | 1.900e-8 | 8.732e-6 | 4.366e-5 | GTP binding | 0.234 | RNA binding | 0.011 |

#### catalytic activity (GO:0003824)

Catalysis of a biochemical reaction at physiological temperatures. In biologically catalyzed reactions, the reactants are known as substrates, and the catalysts are naturally occurring macromolecular substances known as enzymes. Enzymes possess specific binding sites for substrates, and are usually composed wholly or largely of protein, but RNA that has catalytic activity (ribozyme) is often also regarded as enzymatic. In this experiment, the algorithm identified **911** differentially expressed gene(s) out of ALL **5,574** gene(s).

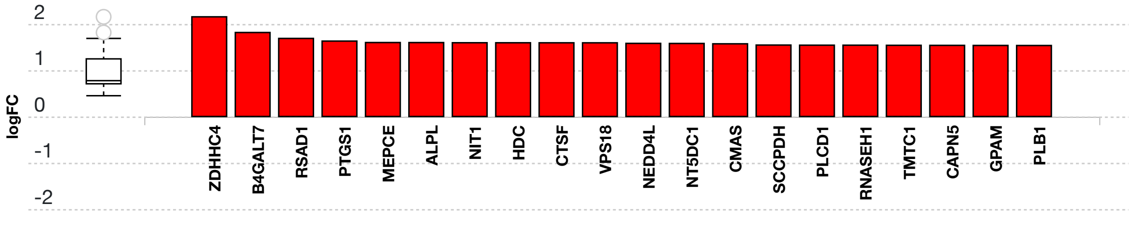


**Fig. 3.3.3: Gene measured expression bar plot:** *All the differentially expressed genes that are annotated to catalytic activity are ranked based on their absolute value of log fold change. The plot is limited to the top 20 genes out of a total of 911 differentially expressed genes. Upregulated genes are shown in red, downregulated genes are shown in blue. The box and whisker plot on the left summarizes the distribution of all the differentially expressed genes that are annotated to this GO term. The box represents the 1st quartile, the median and the 3rd quartile, while the outliers are represented by circles.*

#### protein binding (GO:0005515)

Binding to a protein. In this experiment, the algorithm identified **1,963** differentially expressed gene(s) out of ALL **13,830** gene(s).

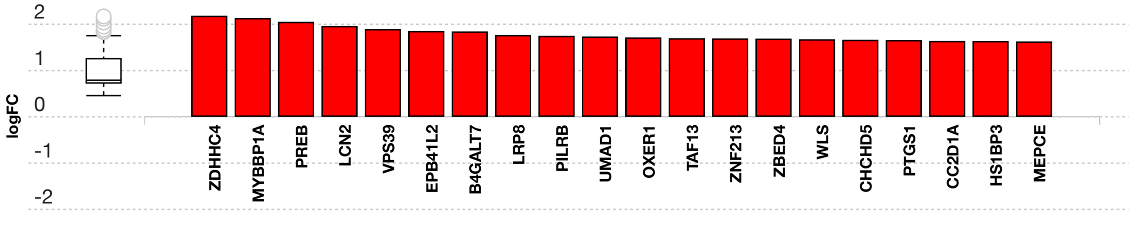

**Fig. 3.3.4: Gene measured expression bar plot:** *All the differentially expressed genes that are annotated to protein binding are ranked based on their absolute value of log fold change. The plot is limited to the top 20 genes out of a total of 1963 differentially expressed genes. Upregulated genes are shown in red, downregulated genes are shown in blue. The box and whisker plot on the left summarizes the distribution of all the differentially expressed genes that are annotated to this GO term. The box represents the 1st quartile, the median and the 3rd quartile, while the outliers are represented by circles.*

### 3.4. Cellular Components results

**Table 3.4.1: Top identified cellular components. Only the top scoring cellular component for each pruning type is described below the table.**

| Pruning Type: None | | | | Pruning Type: High-specificity | | Pruning Type: Smallest Common Denominator | |
|---|---|---|---|---|---|---|---|
| GO Term | p-value | p-value (FDR) | p-value (Bonferroni) | GO Term | p-value | GO Term | p-value |
| intracellular anatomical structure | **1.000e-24** | **1.000e-24** | **1.000e-24** | nucleoplasm | **4.388e-21** | cytoplasm | **1.000e-24** |
| intracellular membrane-bounded organelle | **1.000e-24** | **1.000e-24** | **1.000e-24** | cytosol | **5.546e-20** | nucleoplasm | **7.314e-23** |
| cytoplasm | **1.000e-24** | **1.000e-24** | **1.000e-24** | mitochondrion | 3.169e-6 | organelle envelope | **6.095e-12** |
| intracellular organelle | **1.000e-24** | **1.000e-24** | **1.000e-24** | mitochondrial matrix | 1.219e-5 | intracellular membrane-bounded organelle | **6.400e-6** |
| membrane-bounded organelle | **1.000e-24** | **1.000e-24** | **1.000e-24** | cytoplasm | 1.463e-5 | transferase complex | **4.876e-4** |

#### intracellular anatomical structure (GO:0005622)

A component of a cell contained within (but not including) the plasma membrane. In eukaryotes it includes the nucleus and cytoplasm. In this experiment, the algorithm identified **2,189** differentially expressed gene(s) out of ALL **15,336** gene(s).

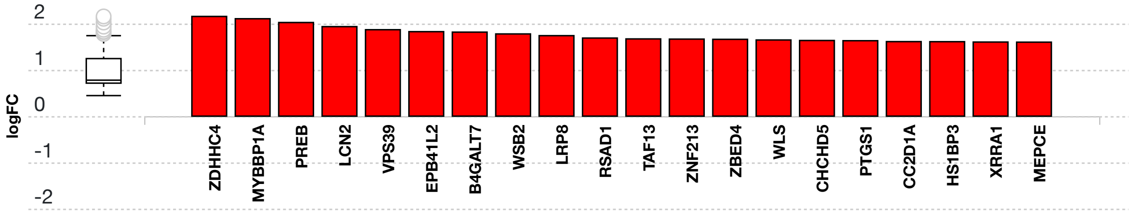


(c) Advaita Corporation 2022

**Fig. 3.4.5: Gene measured expression bar plot:** *All the differentially expressed genes that are annotated to intracellular anatomical structure are ranked based on their absolute value of log fold change. The plot is limited to the top 20 genes out of a total of 2189 differentially expressed genes. Upregulated genes are shown in red, downregulated genes are shown in blue. The box and whisker plot on the left summarizes the distribution of all the differentially expressed genes that are annotated to this GO term. The box represents the 1st quartile, the median and the 3rd quartile, while the outliers are represented by circles.*

#### nucleoplasm (GO:0005654)

That part of the nuclear content other than the chromosomes or the nucleolus. In this experiment, the algorithm identified **720** differentially expressed gene(s) out of ALL **4,085** gene(s).

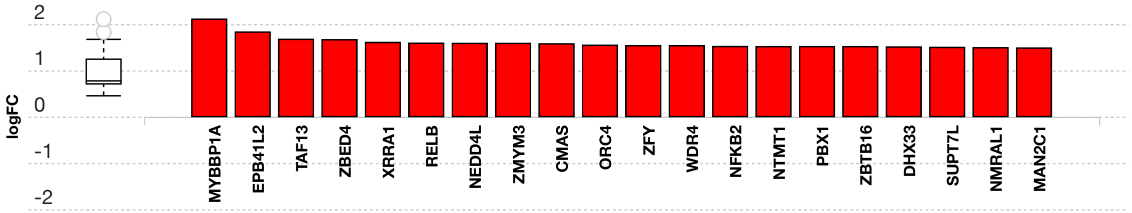


(c) Advaita Corporation 2022

**Fig. 3.4.6: Gene measured expression bar plot:** *All the differentially expressed genes that are annotated to nucleoplasm are ranked based on their absolute value of log fold change. The plot is limited to the top 20 genes out of a total of 720 differentially expressed genes. Upregulated genes are shown in red, downregulated genes are shown in blue. The box and whisker plot on the left summarizes the distribution of all the differentially expressed genes that are annotated to this GO term. The box represents the 1st quartile, the median and the 3rd quartile, while the outliers are represented by circles.*

##### cytoplasm (GO:0005737)

The contents of a cell excluding the plasma membrane and nucleus, but including other subcellular structures. In this experiment, the algorithm identified **1,814** differentially expressed gene(s) out of ALL **11,909** gene(s).

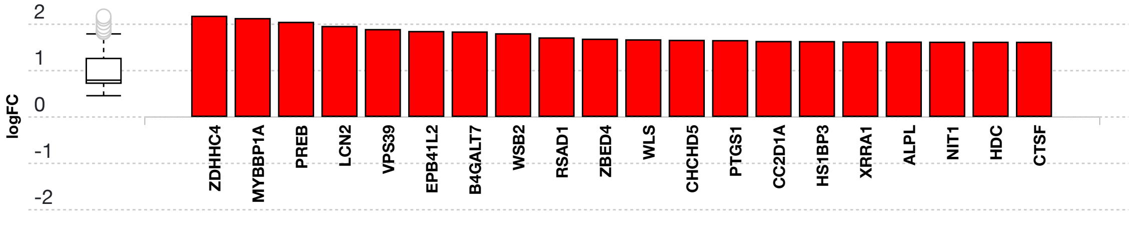


**Fig. 3.4.7: Gene measured expression bar plot:** *All the differentially expressed genes that are annotated to cytoplasm are ranked based on their absolute value of log fold change. The plot is limited to the top 20 genes out of a total of 1814 differentially expressed genes. Upregulated genes are shown in red, downregulated genes are shown in blue. The box and whisker plot on the left summarizes the distribution of all the differentially expressed genes that are annotated to this GO term. The box represents the 1st quartile, the median and the 3rd quartile, while the outliers are represented by circles.*

### 4. Predicted Upstream Regulator Analysis - miRNAs

#### 4.1. Methods

The prediction of active miRNAs (Friedman et al., 2009; Lewis et al., 2005) is based on enrichment of differentially downregulated target genes of the miRNAs. In general, miRNAs have an inhibitory effect on their targets. Therefore, for any given miRNA the method computes the ratio between the number of differentially downregulated targets and all differentially expressed targets, and compares it to the ratio of all downwardly expressed targets to all targets. Overall, iPathwayGuide calculates the probability of observing at least the number of differentially downregulated target genes for a given miRNA just by chance. This p-value is computed using the hypergeometric distribution as described for pORA in the Pathway Analysis section.

#### 4.2. Results

**Table 4.2.1: Top identified miRNAs**

| miRNA Name | p-value | p-value (FDR) | p-value (Bonferroni) |
|---|---|---|---|
| hsa-miR-34c-5p | 1.000 | 1.000 | 1.000 |
| hsa-miR-892c-3p | 1.000 | 1.000 | 1.000 |
| hsa-miR-330-3p | 1.000 | 1.000 | 1.000 |
| hsa-let-7g-5p | 1.000 | 1.000 | 1.000 |
| hsa-miR-299-3p | 1.000 | 1.000 | 1.000 |

#### hsa-miR-34c-5p (MIMAT0000686)

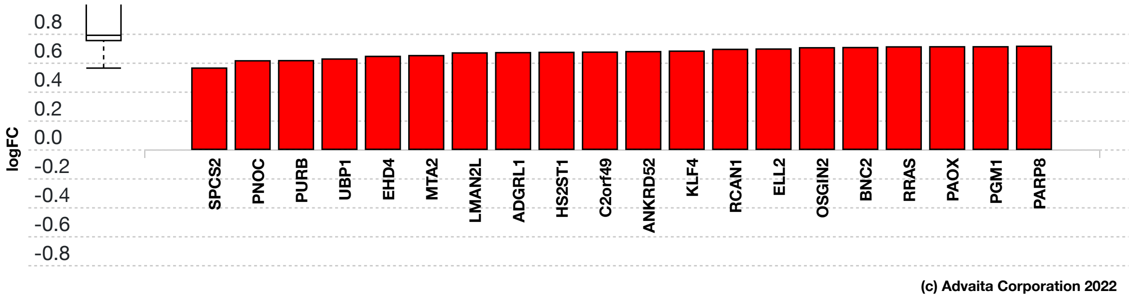


**Fig. 4.2.1: Gene measured expression bar plot:** *All the differentially expressed genes that are targeted by hsa-miR-34c-5p are ranked based on their measured expression change (most downregulated to upregulated). The downregulated genes are shown in blue, and the upregulated ones are shown in red (where applicable). The plot is limited to the top 20 genes out of a total of 103 differentially expressed target genes. Out of all the differentially expressed target genes, 0 were found to be downregulated. The box and whisker plot on the left summarizes the distribution of all the differentially expressed genes targeted by this miRNA. The box represents the 1st quartile, the median and the 3rd quartile, while the outliers are represented by circles.*

#### hsa-miR-892c-3p (MIMAT0025858)

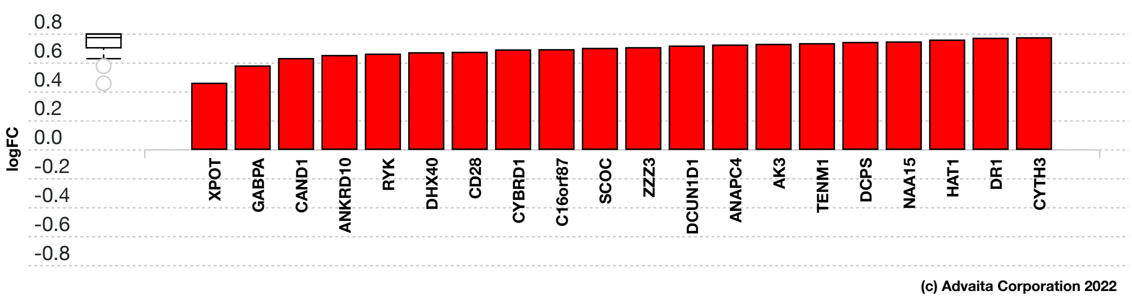


**Fig. 4.2.2: Gene measured expression bar plot:** *All the differentially expressed genes that are targeted by hsa-miR-892c-3p are ranked based on their measured expression change (most downregulated to upregulated). The downregulated genes are shown in blue, and the upregulated ones are shown in red (where applicable). The plot is limited to the top 20 genes out of a total of 41 differentially expressed target genes. Out of all the differentially expressed target genes, 0 were found to be downregulated. The box and whisker plot on the left summarizes the distribution of all the differentially expressed genes targeted by this miRNA. The box represents the 1st quartile, the median and the 3rd quartile, while the outliers are represented by circles.*

#### hsa-miR-330-3p (MIMAT0000751)

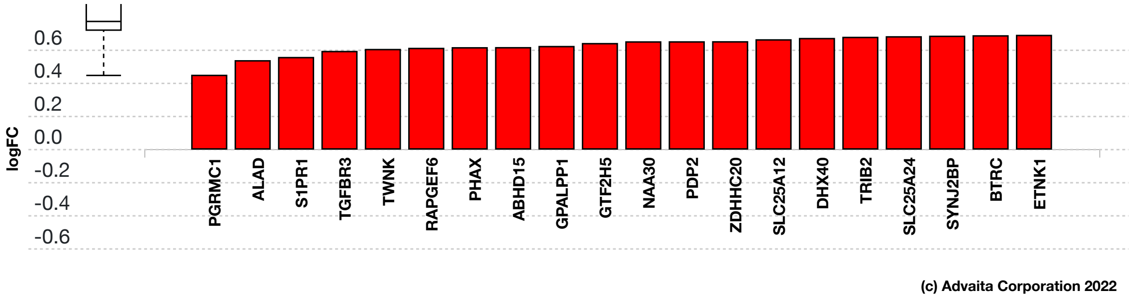


**Fig. 4.2.3: Gene measured expression bar plot:** *All the differentially expressed genes that are targeted by hsa-miR-330-3p are ranked based on their measured expression change (most downregulated to upregulated). The downregulated genes are shown in blue, and the upregulated ones are shown in red (where applicable). The plot is limited to the top 20 genes out of a total of 153 differentially expressed target genes. Out of all the differentially expressed target genes, 0 were found to be downregulated. The box and whisker plot on the left summarizes the distribution of all the differentially expressed genes targeted by this miRNA. The box represents the 1st quartile, the median and the 3rd quartile, while the outliers are represented by circles.*

#### hsa-let-7g-5p (MIMAT0000414)

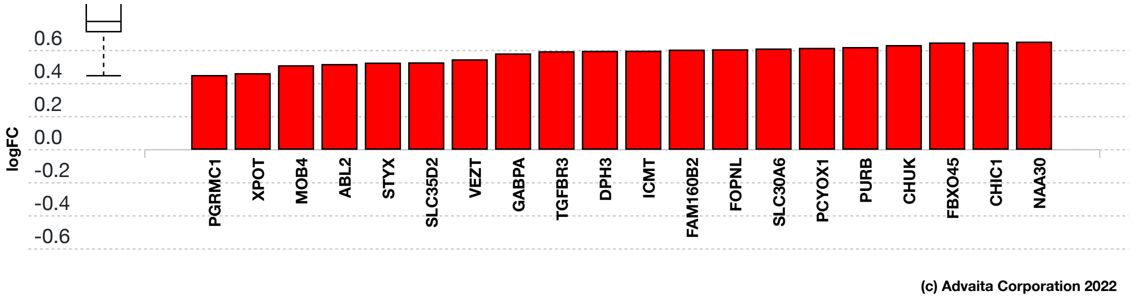


**Fig. 4.2.4: Gene measured expression bar plot:** *All the differentially expressed genes that are targeted by hsa-let-7g-5p are ranked based on their measured expression change (most downregulated to upregulated). The downregulated genes are shown in blue, and the upregulated ones are shown in red (where applicable). The plot is limited to the top 20 genes out of a total of 193 differentially expressed target genes. Out of all the differentially expressed target genes, 0 were found to be downregulated. The box and whisker plot on the left summarizes the distribution of all the differentially expressed genes targeted by this miRNA. The box represents the 1st quartile, the median and the 3rd quartile, while the outliers are represented by circles.*

#### hsa-miR-299-3p (MIMAT0000687)

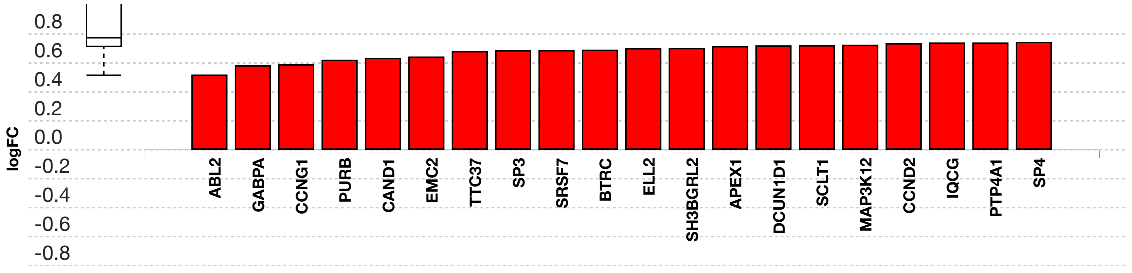


**Fig. 4.2.5: Gene measured expression bar plot:** *All the differentially expressed genes that are targeted by hsa-miR-299-3p are ranked based on their measured expression change (most downregulated to upregulated). The downregulated genes are shown in blue, and the upregulated ones are shown in red (where applicable). The plot is limited to the top 20 genes out of a total of 51 differentially expressed target genes. Out of all the differentially expressed target genes, 0 were found to be downregulated. The box and whisker plot on the left summarizes the distribution of all the differentially expressed genes targeted by this miRNA. The box represents the 1st quartile, the median and the 3rd quartile, while the outliers are represented by circles.*

### 5. Predicted Upstream Regulator Analysis - Genes

#### 5.1. Methods

The prediction of upstream regulators is based on two types of information: i) the enrichment of differentially expressed genes from the experiment and ii) a network of regulatory interactions from our proprietary knowledge base (see the report information for details). The network is a directed graph in which the nodes represent genes, and the edges represent regulatory interactions between two genes. A signed edge in this graph consists of a source gene, a target gene, and a sign to indicate the type of signal: activation (+) or inhibition (-). To create the network, the analysis selects only those edges observed in the literature with at least a medium confidence (evidence score greater than or equal to 400). The analysis considers two hypotheses:

HA. The upstream regulator is **activated** in the condition studied.
HI. The upstream regulator is **inhibited** in the condition studied.

The analysis divides the set of all the genes obtained from NCBI Gene database into several subsets based on the measurements in the experiment and the definitions shown in **Figure 5.1.1** and **Figure 5.1.2**. Let the sign of a measured DE gene be the sign of the log fold change value: (+) for up-regulated genes and (-) for down-regulated genes. A gene is a target gene if it corresponds to a node in the network that has at least one incoming edge. We define a *consistent gene* as a target DE gene such that the sign of the gene is consistent both with the type of the signal **and** with the hypothesis considered. Formally, by definition, a target DE gene *g* is consistent with Hypothesis HA if and only if an incoming edge *e* exists such that *sign(g) = sign(e)*. In other words, this describes the situation when the upstream regulator is predicted as activated, the signal is activation and the target DE gene is up-regulated, or the signal is inhibition and the target DE gene is down-regulated (see panel A in **Figure 5.1.1**). A target DE gene *g* is consistent with Hypothesis HI if and only if an incoming edge *e* exists such that *sign(g) ≠ sign(e)*. This second case captures the situation in which the upstream regulator is inhibited, the signal is inhibition and the target DE gene is up-regulated, or the signal is activation and the target DE gene is down-regulated (see panel B in **Figure 5.1.1**).

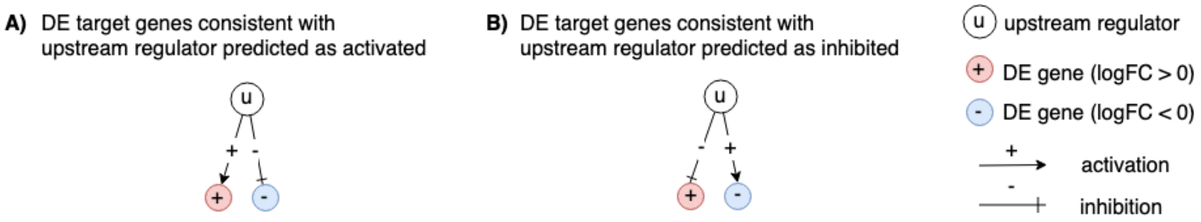


**Fig. 5.1.1: Target genes consistent with the hypothesis considered:** *In panel A, the signs of the DE genes match the signs of their respective incoming edges, increasing the likelihood that the upstream regulator u is activated. In panel B, the signs of the DE genes are opposite to the signs of their edges, increasing the likelihood that the upstream regulator u is inhibited.*

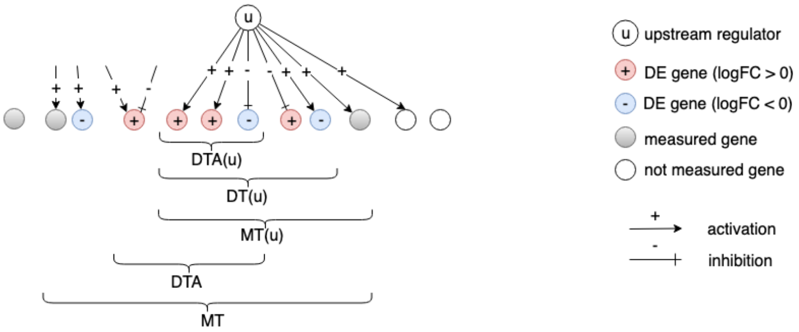


**Fig. 5.1.2:** *The set of all genes includes the set of measured genes that are also targets in the network, or Measured Targets (MT). We define the subset of "DE Targets consistent with the first hypothesis that the upstream regulators are Activated", DTA. For a selected upstream regulator u, we have the set of "Measured Targets of u" MT(u), "Differentially expressed Targets downstream of u" DT(u), and the set of "DE targets consistent with the hypothesis HA that u is Activated" DTA(u). The equivalent graphic for the hypothesis HI associated with DTI and DTI(u) is*

*not shown.*

#### Upstream regulators Z-score

For both research hypotheses, the analysis computes a Z-score for each upstream regulator *z(u)* by iterating over the genes in DT(u) and their incoming edges *in(g)*. We can then compute the p-value corresponding to the z-score $P_z$ as the one-tailed area under the probability density function for a normal distribution, N(0,1).

#### Upstream regulators predicted as activated

Here, the research hypothesis considers the upstream regulator as activated. For each upstream regulator *u*, the number of consistent DE genes downstream of *u*, DTA(u) is compared to the number of measured target genes expected to be both consistent and DE just by chance. iPathwayGuide uses an over-representation approach to compute the statistical significance of observing at least the given number of consistent DE genes. The p-value $P_{act}$ is computed using the hypergeometric distribution (Draghici et al., 2003, Draghici 2011).

After computing a p-value for both types of evidence, $P_z$ and $P_{act}$, we need to combine these two probabilities into one global probability value, $P_G$ that is used to rank the upstream regulators and test the research hypothesis that the upstream regulators are predicted as activated in the condition studied. Since only a positive z-score indicates that the upstream regulator is predicted as activated, we only combine p-values for a positive z-score. Moreover, to avoid introducing false positives, only $P_z$ for significant z-scores ( $z \geq 2$ ) are combined. The analysis uses the standard Fisher's method to combine p-values into one test statistic (Fisher 1925).

#### Upstream regulators predicted as inhibited

In parallel with upstream regulators predicted as activated, we use $P_{inh}$ and $P_z$ to predict upstream regulators that are inhibited. Here, the research hypothesis states that the upstream regulators are inhibited in the conditions studied. For each upstream regulator *u*, the number of consistent DE genes downstream of *u*, DTI(u) is compared to the number of measured target genes expected to be both consistent and DE just by chance. Using the Fisher's method as above, the analysis combines $P_{inh}$ and $P_z$, where $P_z$ is considered only for significant negative z-scores ( $z \leq -2$).

### 5.2. Results: upstream regulators predicted as activated

| Upstream Regulator (u) | DTA(u) | DT(u) | p-value | p-value (FDR) | p-value (Bonferroni) |
|---|---|---|---|---|---|
| RANBP2 | 38 | 38 | **9.188e-14** | **1.664e-10** | **1.664e-10** |
| NUP160 | 36 | 36 | **6.001e-13** | **4.112e-10** | **1.087e-9** |
| NUP107 | 36 | 36 | **6.811e-13** | **4.112e-10** | **1.233e-9** |
| NUP43 | 35 | 35 | **1.624e-12** | **6.657e-10** | **2.941e-9** |
| NUP37 | 35 | 35 | **1.838e-12** | **6.657e-10** | **3.328e-9** |

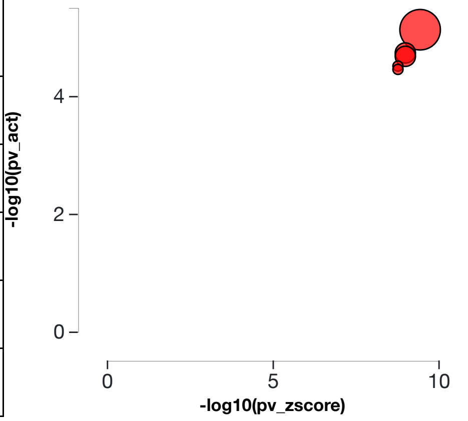


**Table 5.2.1: Top upstream regulators predicted as activated.** *For each upstream regulator u, the table shows the number of DE targets supporting the hypothesis that the regulator is activated DTA(u) the total number of DE genes downstream of u DT(u), the combined raw p-value, and the p-value corrected for multiple comparisons.* **Fig. 5.2.1: A two-way plot showing the top five upstream regulators predicted as activated.** *Dots representing upstream regulators are positioned using $P_{zscore}$ on the horizontal axis, and using $P_{act}$ on the vertical axis. $P_{act}$ is the p-value based on the number of DE targets consistent with the typeof the incoming signal and with the selected hypothesis type. Upstream regulators with a significant combined p-value are shown in red. The size of each dot represents the number of consistent DE genes for that regulator.*

#### RANBP2 (RAN binding protein 2)

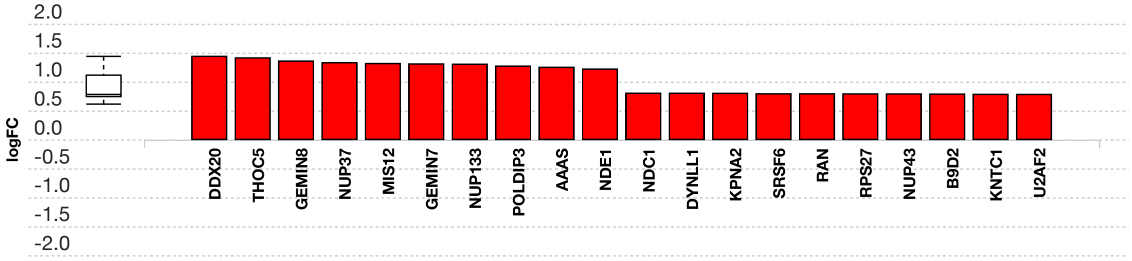


**Fig. 5.2.3: Gene measured expression bar plot:** *All the consistent differentially expressed genes that are targeted by RANBP2 are ranked based on their absolute value of log fold change. The plot is limited to the top 20 genes out of a total of 38 consistent differentially expressed target genes. Upregulated genes are shown in red, downregulated genes are shown in blue. The box and whisker plot on the left summarizes the distribution of all the consistent differentially expressed genes targeted by this upstream regulator. The box shows the 1st quartile, the median and the 3rd quartile, while the outliers are represented by circles.*

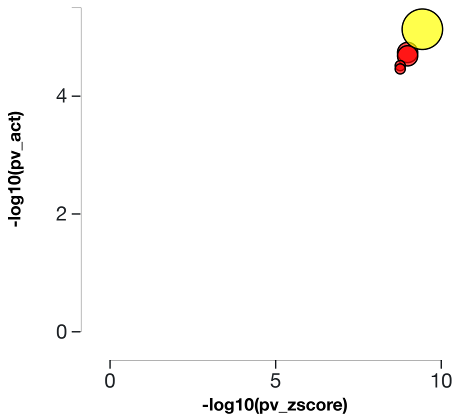


**Fig. 5.2.4: Activation p-value vs zscore p-value:** *RANBP2, RAN binding protein 2, (yellow) is shown, using negative log of the activation and zscore p-values, along with the other most significant upstream regulators. The size of the dot represents the relative number of consistent DE genes, which for selected upstream regulator is 38.*

#### NUP160 (nucleoporin 160)

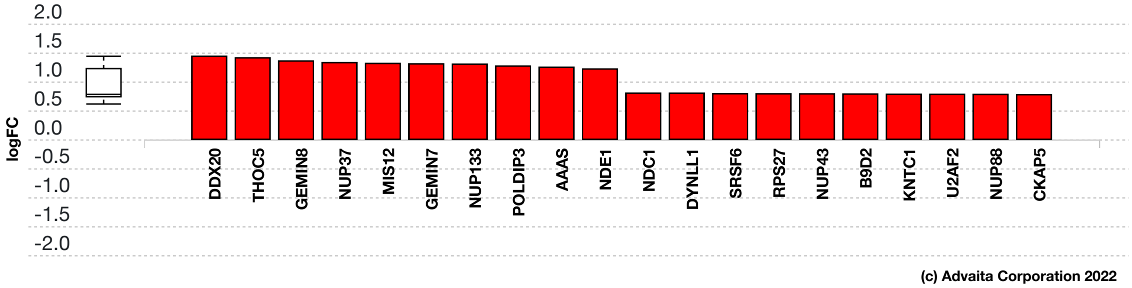


**Fig. 5.2.5: Gene measured expression bar plot:** *All the consistent differentially expressed genes that are targeted by NUP160 are ranked based on their absolute value of log fold change. The plot is limited to the top 20 genes out of a total of 36 consistent differentially expressed target genes. Upregulated genes are shown in red, downregulated genes are shown in blue. The box and whisker plot on the left summarizes the distribution of all the consistent differentially expressed genes targeted by this upstream regulator. The box shows the 1st quartile, the median and the 3rd quartile, while the outliers are represented by circles.*

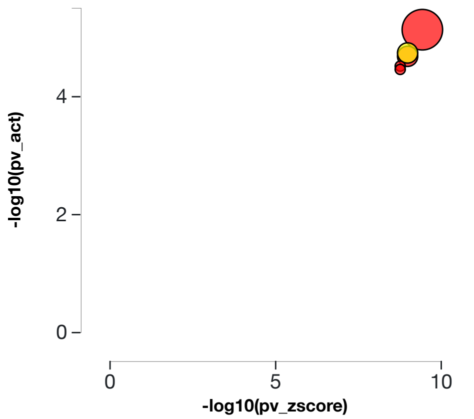


**Fig. 5.2.6: Activation p-value vs zscore p-value:** *NUP160, nucleoporin 160, (yellow) is shown, using negative log of the activation and zscore p-values, along with the other most significant upstream regulators. The size of the dot represents the relative number of consistent DE genes, which for selected upstream regulator is 36.*

#### NUP107 (nucleoporin 107)

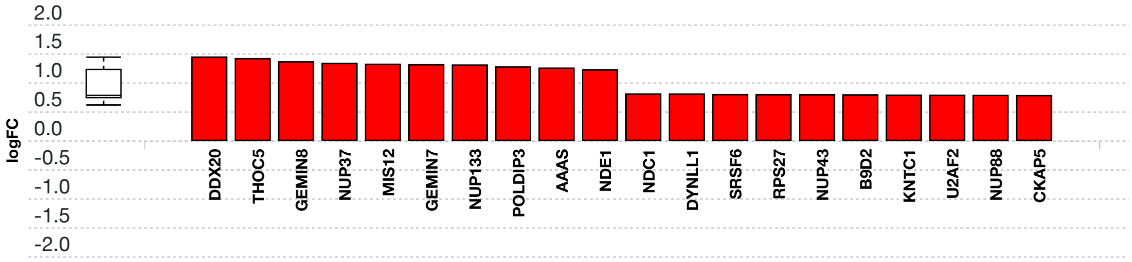


**Fig. 5.2.7: Gene measured expression bar plot:** *All the consistent differentially expressed genes that are targeted by NUP107 are ranked based on their absolute value of log fold change. The plot is limited to the top 20 genes out of a total of 36 consistent differentially expressed target genes. Upregulated genes are shown in red, downregulated genes are shown in blue. The box and whisker plot on the left summarizes the distribution of all the consistent differentially expressed genes targeted by this upstream regulator. The box shows the 1st quartile, the median and the 3rd quartile, while the outliers are represented by circles.*

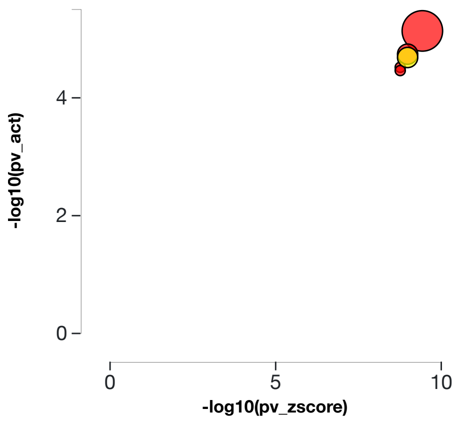


**Fig. 5.2.8: Activation p-value vs zscore p-value:** *NUP107, nucleoporin 107, (yellow) is shown, using negative log of the activation and zscore p-values, along with the other most significant upstream regulators. The size of the dot represents the relative number of consistent DE genes, which for selected upstream regulator is 36.*

#### NUP43 (nucleoporin 43)

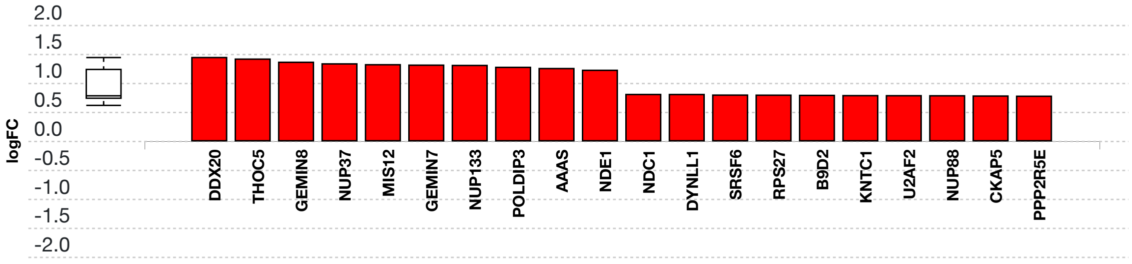


**Fig. 5.2.9: Gene measured expression bar plot:** *All the consistent differentially expressed genes that are targeted by NUP43 are ranked based on their absolute value of log fold change. The plot is limited to the top 20 genes out of a total of 35 consistent differentially expressed target genes. Upregulated genes are shown in red, downregulated genes are shown in blue. The box and whisker plot on the left summarizes the distribution of all the consistent differentially expressed genes targeted by this upstream regulator. The box shows the 1st quartile, the median and the 3rd quartile, while the outliers are represented by circles.*

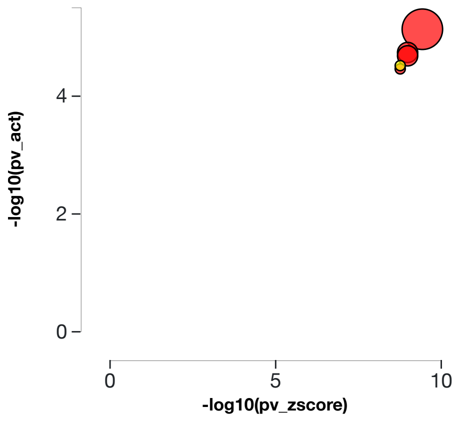


**Fig. 5.2.10: Activation p-value vs zscore p-value:** *NUP43, nucleoporin 43, (yellow) is shown, using negative log of the activation and zscore p-values, along with the other most significant upstream regulators. The size of the dot represents the relative number of consistent DE genes, which for selected upstream regulator is 35.*

#### NUP37 (nucleoporin 37)

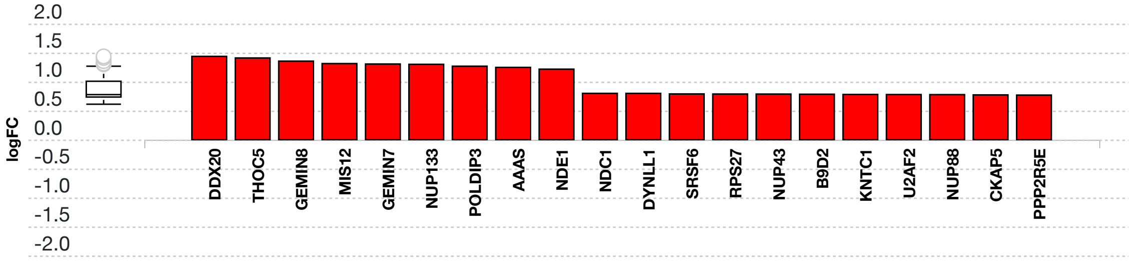


**Fig. 5.2.11: Gene measured expression bar plot:** *All the consistent differentially expressed genes that are targeted by NUP37 are ranked based on their absolute value of log fold change. The plot is limited to the top 20 genes out of a total of 35 consistent differentially expressed target genes. Upregulated genes are shown in red, downregulated genes are shown in blue. The box and whisker plot on the left summarizes the distribution of all the consistent differentially expressed genes targeted by this upstream regulator. The box shows the 1st quartile, the median and the 3rd quartile, while the outliers are represented by circles.*

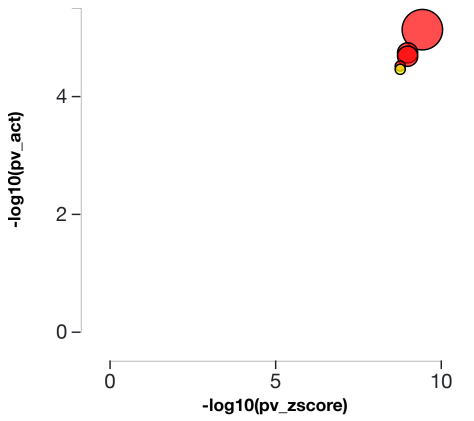


**Fig. 5.2.12: Activation p-value vs zscore p-value:** *NUP37, nucleoporin 37, (yellow) is shown, using negative log of the activation and zscore p-values, along with the other most significant upstream regulators. The size of the dot represents the relative number of consistent DE genes, which for selected upstream regulator is 35.*

### 5.3. Results: upstream regulators predicted as inhibited

| Upstream Regulator (u) | DTI(u) | DT(u) | p-value | p-value (FDR) | p-value (Bonferroni) |
|---|---|---|---|---|---|
| RBX1 | 19 | 19 | 1.077e-11 | 4.952e-9 | 1.950e-8 |
| SKP2 | 17 | 17 | 2.189e-11 | 4.952e-9 | 3.965e-8 |
| COMMD3 | 18 | 18 | 2.415e-11 | 4.952e-9 | 4.374e-8 |
| CCDC22 | 18 | 18 | 2.415e-11 | 4.952e-9 | 4.374e-8 |
| COMMD2 | 18 | 18 | 2.415e-11 | 4.952e-9 | 4.374e-8 |

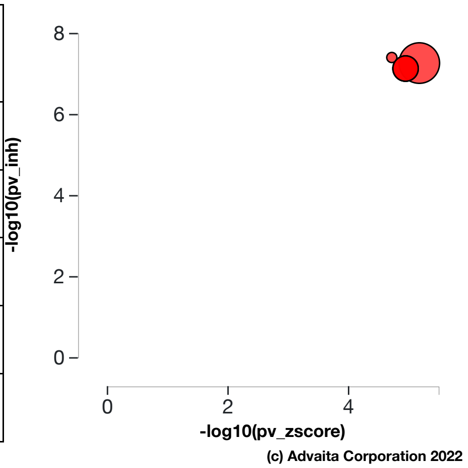


**Table 5.3.1: Top upstream regulators predicted as inhibited.** *For each upstream regulator u, the table shows the number of DE targets supporting the hypothesis that the regulator is inhibited DTI(u) the total number of DE genes downstream of u DT(u), the combined raw p-value, and the p-value corrected for multiple comparisons.* **Fig. 5.3.1: A two-way plot showing the top five upstream regulators predicted as inhibited.** *Dots representing upstream regulators are positioned using $P_{zscore}$ on the horizontal axis, and using $P_{inh}$ on the vertical axis. $P_{inh}$ is the p-value based on the number of DE targets consistent with the typeof the incoming signal and with the selected hypothesis type. Upstream regulators with a significant combined p-value are shown in red. The size of each dot represents the number of consistent DE genes for that regulator.*

#### RBX1 (ring-box 1)

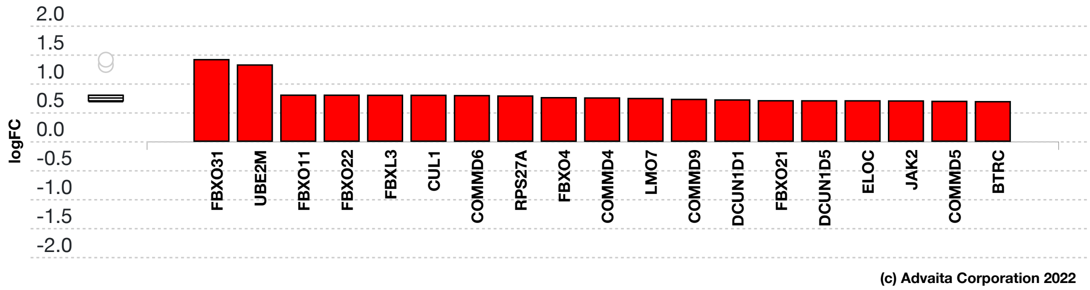


**Fig. 5.3.13: Gene measured expression bar plot:** *All the consistent differentially expressed genes that are targeted by RBX1 are ranked based on their absolute value of log fold change. Upregulated genes are shown in red, downregulated genes are shown in blue. The box and whisker plot on the left summarizes the distribution of all the consistent differentially expressed genes targeted by this upstream regulator. The box shows the 1st quartile, the median and the 3rd quartile, while the outliers are represented by circles.*

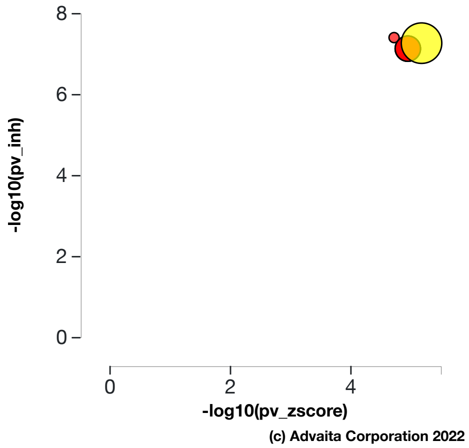


**Fig. 5.3.14: Inhibition p-value vs zscore p-value:** *RBX1, ring-box 1, (yellow) is shown, using negative log of the inhibition and zscore p-values, along with the other most significant upstream regulators. The size of the dot represents the relative number of consistent DE genes, which for selected upstream regulator is 19.*

#### SKP2 (S-phase kinase associated protein 2)

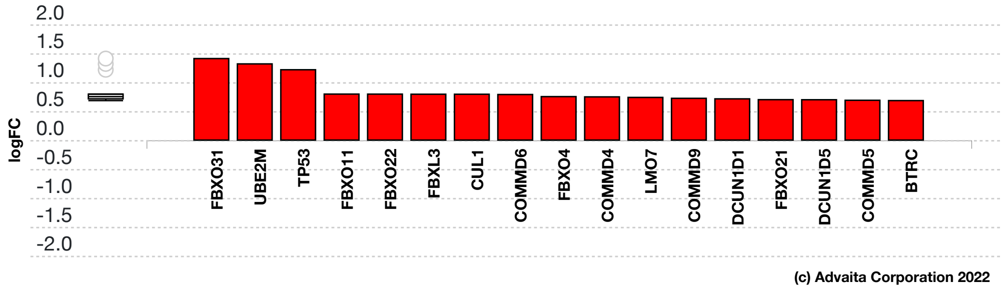


**Fig. 5.3.15: Gene measured expression bar plot:** *All the consistent differentially expressed genes that are targeted by SKP2 are ranked based on their absolute value of log fold change. Upregulated genes are shown in red, downregulated genes are shown in blue. The box and whisker plot on the left summarizes the distribution of all the consistent differentially expressed genes targeted by this upstream regulator. The box shows the 1st quartile, the median and the 3rd quartile, while the outliers are represented by circles.*

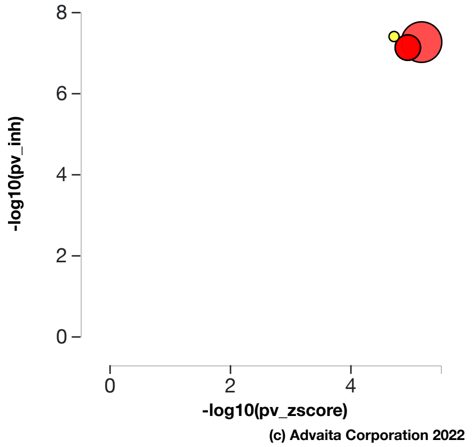


**Fig. 5.3.16: Inhibition p-value vs zscore p-value:** *SKP2, S-phase kinase associated protein 2, (yellow) is shown, using negative log of the inhibition and zscore p-values, along with the other most significant upstream regulators. The size of the dot represents the relative number of consistent DE genes, which for selected upstream regulator is 17.*

#### COMMD3 (COMM domain containing 3)

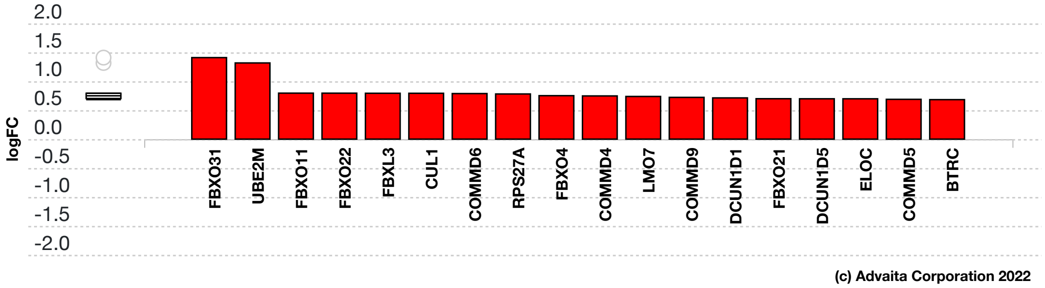


**Fig. 5.3.17: Gene measured expression bar plot:** *All the consistent differentially expressed genes that are targeted by COMMD3 are ranked based on their absolute value of log fold change. Upregulated genes are shown in red, downregulated genes are shown in blue. The box and whisker plot on the left summarizes the distribution of all the consistent differentially expressed genes targeted by this upstream regulator. The box shows the 1st quartile, the median and the 3rd quartile, while the outliers are represented by circles.*

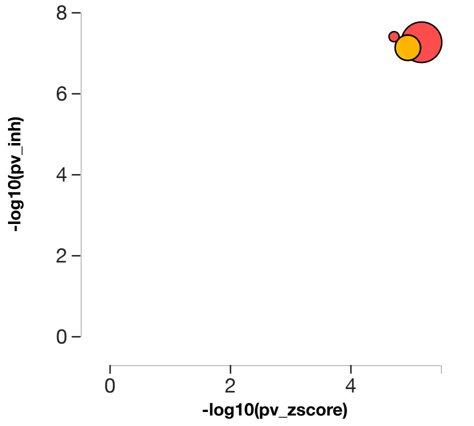


**Fig. 5.3.18: Inhibition p-value vs zscore p-value:** *COMMD3, COMM domain containing 3, (yellow) is shown, using negative log of the inhibition and zscore p-values, along with the other most significant upstream regulators. The size of the dot represents the relative number of consistent DE genes, which for selected upstream regulator is 18.*

#### CCDC22 (coiled-coil domain containing 22)

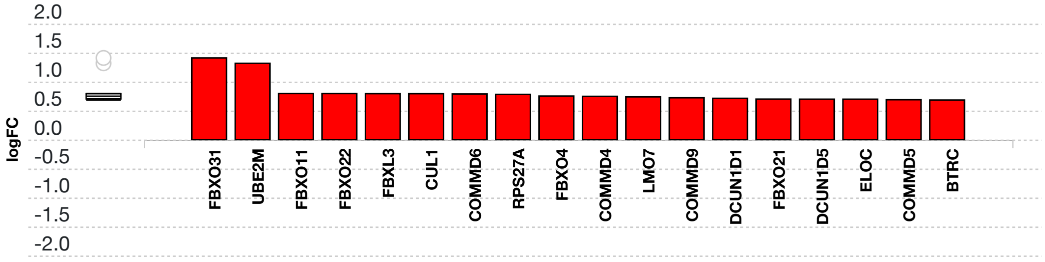


**Fig. 5.3.19: Gene measured expression bar plot:** *All the consistent differentially expressed genes that are targeted by CCDC22 are ranked based on their absolute value of log fold change. Upregulated genes are shown in red, downregulated genes are shown in blue. The box and whisker plot on the left summarizes the distribution of all the consistent differentially expressed genes targeted by this upstream regulator. The box shows the 1st quartile, the median and the 3rd quartile, while the outliers are represented by circles.*

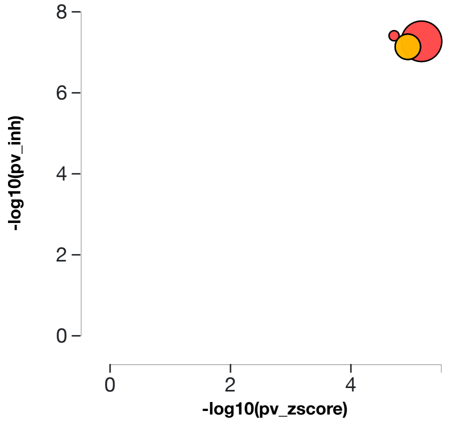


**Fig. 5.3.20: Inhibition p-value vs zscore p-value:** *CCDC22, coiled-coil domain containing 22, (yellow) is shown, using negative log of the inhibition and zscore p-values, along with the other most significant upstream regulators. The size of the dot represents the relative number of consistent DE genes, which for selected upstream regulator is 18.*

#### COMMD2 (COMM domain containing 2)

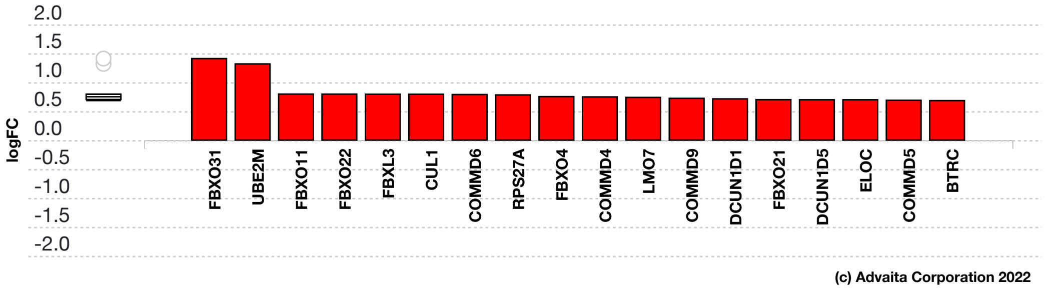


**Fig. 5.3.21: Gene measured expression bar plot:** *All the consistent differentially expressed genes that are targeted by COMMD2 are ranked based on their absolute value of log fold change. Upregulated genes are shown in red, downregulated genes are shown in blue. The box and whisker plot on the left summarizes the distribution of all the consistent differentially expressed genes targeted by this upstream regulator. The box shows the 1st quartile, the median and the 3rd quartile, while the outliers are represented by circles.*

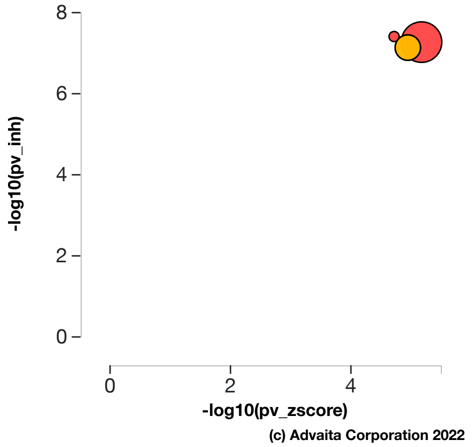


**Fig. 5.3.22: Inhibition p-value vs zscore p-value:** *COMMD2, COMM domain containing 2, (yellow) is shown, using negative log of the inhibition and zscore p-values, along with the other most significant upstream regulators. The size of the dot represents the relative number of consistent DE genes, which for selected upstream regulator is 18.*

### 6. Predicted Upstream Regulator Analysis – Chemicals, Drugs, Toxicants (CDTs)

#### 6.1. Methods

The prediction of upstream Chemicals, Drugs, Toxicants (CDTs) is based on two types of information: i) the enrichment of differentially expressed genes from the experiment and ii) a network of interactions from the Advaita Knowledge Base (AKB v2201). The network is a directed graph in which the source node represents either a chemical substance or compound (e.g. zinc), a drug (e.g. aspirin), or a toxicant (e.g. tobacco smoke). The generic abbreviation CDT will be used henceforth to designate any of these. The edges represent known effects that these CDTs have on various genes. A signed edge in this graph consists of a source CDT, a target gene, and a sign to indicate the type of effect: activation (+) or inhibition (-). The analysis considers two hypotheses:

HP. The upstream chemical, drug or toxicant is **present (or overly abundant)** in the condition studied.
HA. The upstream chemical, drug or toxicant is **absent (or insufficient)** in the condition studied.

The analysis divides the set of all the genes from AKB into several subsets based on the measurements in the experiment and the definitions shown in **Figure 6.1.1** and **Figure 6.1.2**. Let the sign of a measured DE gene be the sign of the log fold change value: (+) for up-regulated genes and (-) for down-regulated genes. A gene is a target gene if it corresponds to a node in the network that has at least one incoming edge. We define a *consistent gene* as a target DE gene such that the sign of the gene is consistent both with the type of the signal **and** with the hypothesis considered. Formally, by definition, a target DE gene $g$ is consistent with Hypothesis HP if and only if an incoming edge $e$ exists such that $sign(g) = sign(e)$. In other words, this describes the situation when the CDT is predicted as present, the signal is activation and the target DE gene is up-regulated, or the signal is inhibition and the target DE gene is down-regulated (see panel A in **Figure 6.1.1**). A target DE gene $g$ is consistent with Hypothesis HA if and only if an incoming edge $e$ exists such that $sign(g) \neq sign(e)$. This second case captures the situation in which the CDT is absent (or insufficient), the signal is inhibition and the target DE gene is up-regulated, or the signal is activation and the target DE gene is down-regulated (see panel B in **Figure 6.1.1**).

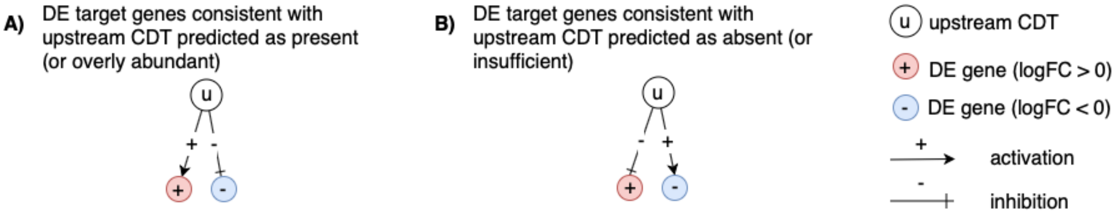


**Fig. 6.1.1: Target genes consistent with the hypothesis considered:** *In panel A, the signs of the DE genes match the signs of their respective incoming edges, increasing the likelihood that the CDT u is present. In panel B, the signs of the DE genes are opposite to the signs of their edges, increasing the likelihood that the CDT u is absent.*

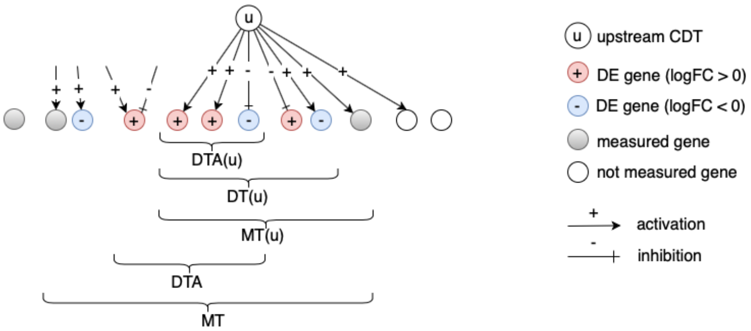


**Fig. 6.1.2:** *The set of all genes includes the set of measured genes that are also targets in the network, or Measured Targets (MT). We define the subset of "DE Targets consistent with the first hypothesis that the CDTs are Present (or overly abundant)", DTA. For a selected upstream CDT u, we have the set of "Measured Targets of u" MT(u), "Differentially expressed Targets downstream of u" DT(u), and the set of "DE targets consistent with the hypothesis HP that u is Present" DTA(u). The equivalent graphic for the hypothesis HA associated with DTI and DTI(u) is not shown.*

#### Z-score

For both research hypotheses, the analysis computes a Z-score for each CDT *z(u)* by iterating over the genes in DT(u) and their incoming edges *in(g)*. We can then compute the p-value corresponding to the z-score $P_z$ as the one-tailed area under the probability density function for a normal distribution, N(0,1).

#### Upstream CDTs predicted as present (or overly abundant)

Here, the research hypothesis considers presence of the CDT. This hypothesis is useful when investigating whether the given phenotype has been impacted by the presence of a given chemical, drug or toxicant (e.g. tobacco smoke, dioxin, etc.). For each CDT *u*, the number of consistent DE genes downstream of *u*, DTA(u) is compared to the number of measured target genes expected to be both consistent and DE just by chance. iPathwayGuide uses an over-representation approach to compute the statistical significance of observing at least the given number of consistent DE genes. The p-value $P_{pres}$ is computed using the hypergeometric distribution (Draghici et al., 2003, Draghici 2011).

After computing a p-value for both types of evidence, $P_z$ and $P_{pres}$, we combine these two probabilities into one global probability value, $P_G$ that is used to rank the upstream regulators and test the research hypothesis that the upstream CDTs are predicted as present in the condition studied. The analysis uses the standard Fisher's method to combine p-values into one test statistic (Fisher 1925).

#### Upstream CDTs predicted as absent (or insufficient)

In parallel with upstream CDTs predicted as present, we use $P_{abs}$ and $P_z$ to predict upstream CDTs that are absent. This hypothesis is relevant when investigating whether the given phenotype has been impacted by the lack of a given chemical that is necessary for the well-functioning of the organism or cell (e.g. a vitamin deficiency, iron deficiency, etc.). Here, the research hypothesis states that the upstream CDT are insufficient in the condition studied. For each upstream CDT *u*, the number of consistent DE genes downstream of *u*, DTI(u) is compared to the number of measured target genes expected to be both consistent and DE just by chance. Using the Fisher's method as above, the analysis combines $P_{abs}$ and $P_z$, where $P_z$ is considered only for significant negative z-scores ( $z \leq -2$).

#### 6.2. Results: upstream CDTs predicted as present (or overly abundant)

| CDT (u) | DTA(u) | DT(u) | p-value | p-value (FDR) | p-value (Bonferroni) |
|---|---|---|---|---|---|
| Naphthoquinones | 62 | 63 | 1.957e-16 | 2.972e-13 | 4.323e-13 |
| geldanamycin | 59 | 61 | 2.437e-15 | 1.794e-12 | 5.383e-12 |
| Dihydrotestosterone | 131 | 135 | 2.691e-16 | 2.972e-13 | 5.944e-13 |
| cylindrospermopsin | 77 | 85 | 1.435e-14 | 7.922e-12 | 3.169e-11 |
| Sodium Selenite | 167 | 224 | 7.077e-13 | 3.127e-10 | 1.563e-9 |

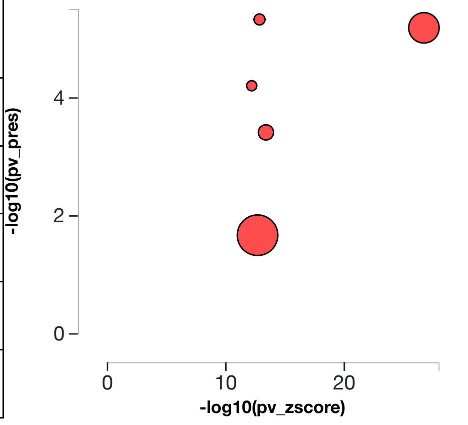


**Table 6.2.1: Top upstream CDTs predicted as present (or overly abundant).** *For each upstream CDT u, the table shows the number of DE targets supporting the hypothesis that the CDT is present DTA(u) the total number of DE genes downstream of u DT(u), the combined raw p-value, and the p-value corrected for multiple comparisons.* **Fig. 6.2.1: A two-way plot showing the top five upstream CDTs predicted as present (or overly abundant).** *Dots representing upstream CDTs are positioned using $P_{zscore}$ on the horizontal axis, and using $P_{pres}$ on the vertical axis. $P_{pres}$ is the p-value based on the number of DE targets consistent with the typeof the incoming signal and with the selected hypothesis type. Upstream CDTs with a significant combined p-value are shown in red. The size of each dot represents the relative number of consistent DE genes for that CDT.*

##### Naphthoquinones

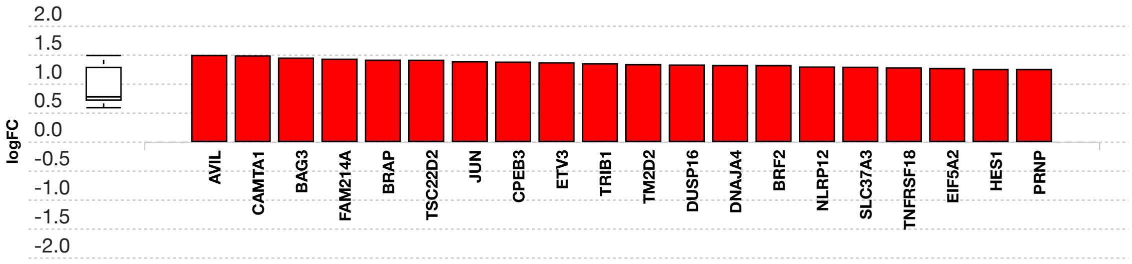


**Fig. 6.2.3: Consistent DE target genes measured expression bar plot:** *All the consistent differentially expressed genes that are targeted by Naphthoquinones are ranked based on their absolute value of log fold change. The plot is limited to the top 20 genes out of a total of 62 consistent differentially expressed target genes. Upregulated genes are shown in red, downregulated genes are shown in blue. The box and whisker plot on the left summarizes the distribution of all the consistent differentially expressed genes targeted by this upstream regulator. The box shows the 1st quartile, the median and the 3rd quartile, while any outliers are represented by circles.*

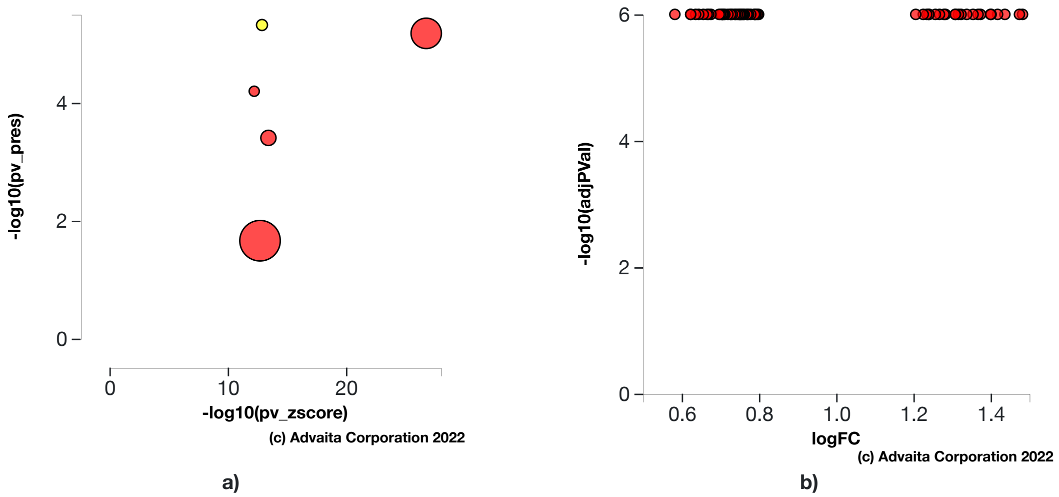


**Fig. 6.2.4: a) Present (overly abundant) p-value vs zscore p-value:** *The significance of Naphthoquinones is plotted on two axes, with negative log of $P_z$ on x-axis and negative log of $P_{pres}$ on y-axis. The size of the dot represents the relative number of consistent DE genes, which for selected upstream regulator is 62.* **b) Volcano plot:** *There are* **62** *DE genes that are targets of Naphthoquinones consistent with the hypothesis that Naphthoquinones is present (overly abundant) The target genes are represented in terms of their measured expression change (x-axis) and the significance of the change (y-axis). The significance is represented in terms of the negative log (base 10) of the p-value, so that more significant genes are plotted higher on the y-axis.*

#### geldanamycin

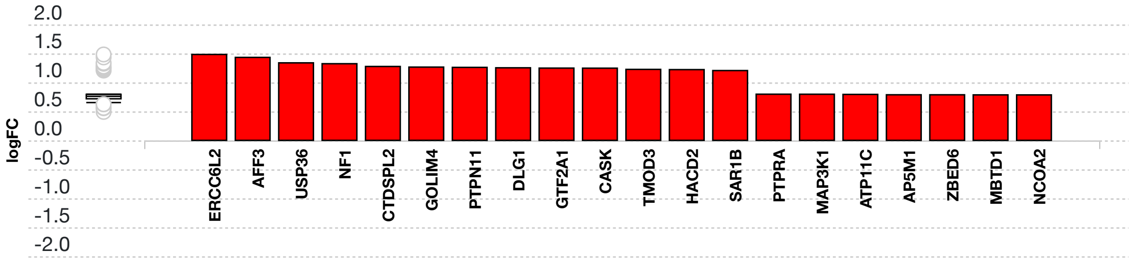


**Fig. 6.2.5: Consistent DE target genes measured expression bar plot:** *All the consistent differentially expressed genes that are targeted by geldanamycin are ranked based on their absolute value of log fold change. The plot is limited to the top 20 genes out of a total of 59 consistent differentially expressed target genes. Upregulated genes are shown in red, downregulated genes are shown in blue. The box and whisker plot on the left summarizes the distribution of all the consistent differentially expressed genes targeted by this upstream regulator. The box shows the 1st quartile, the median and the 3rd quartile, while any outliers are represented by circles.*

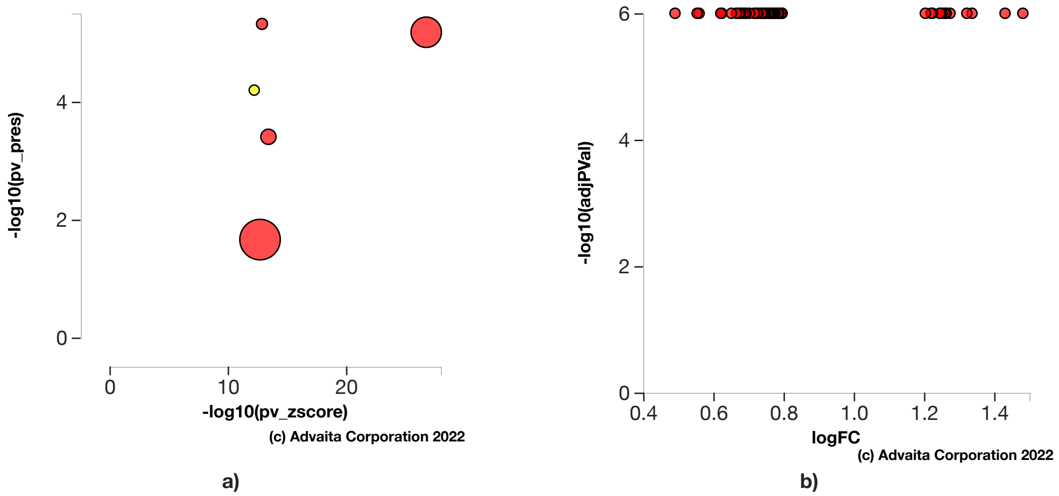


**Fig. 6.2.6: a) Present (overly abundant) p-value vs zscore p-value:** *The significance of geldanamycin is plotted on two axes, with negative log of $P_z$ on x-axis and negative log of $P_{pres}$ on y-axis. The size of the dot represents the relative number of consistent DE genes, which for selected upstream regulator is 59.* **b) Volcano plot:** *There are **59** DE genes that are targets of geldanamycin consistent with the hypothesis that geldanamycin is present (overly abundant) The target genes are represented in terms of their measured expression change (x-axis) and the significance of the change (y-axis). The significance is represented in terms of the negative log (base 10) of the p-value, so that more significant genes are plotted higher on the y-axis.*

#### Dihydrotestosterone

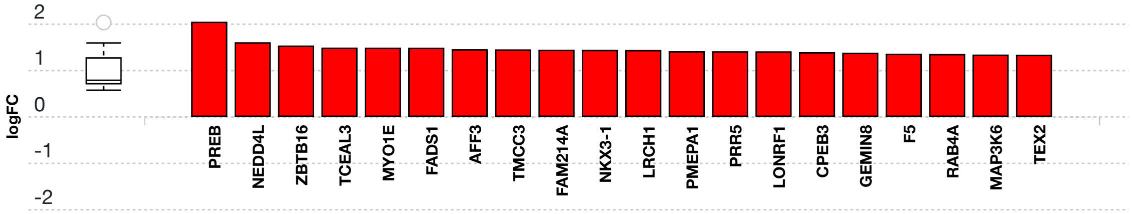


**Fig. 6.2.7: Consistent DE target genes measured expression bar plot:** *All the consistent differentially expressed genes that are targeted by Dihydrotestosterone are ranked based on their absolute value of log fold change. The plot is limited to the top 20 genes out of a total of 131 consistent differentially expressed target genes. Upregulated genes are shown in red, downregulated genes are shown in blue. The box and whisker plot on the left summarizes the distribution of all the consistent differentially expressed genes targeted by this upstream regulator. The box shows the 1st quartile, the median and the 3rd quartile, while any outliers are represented by circles.*

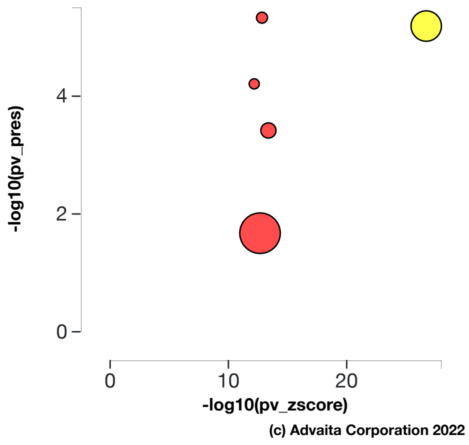


a)

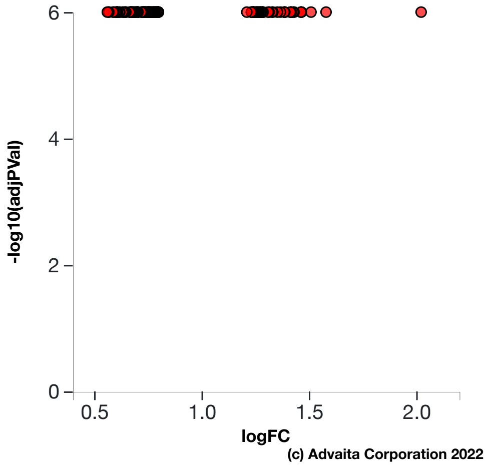


b)

**Fig. 6.2.8: a) Present (overly abundant) p-value vs zscore p-value:** *The significance of Dihydrotestosterone is plotted on two axes, with negative log of $P_z$ on x-axis and negative log of $P_{pres}$ on y-axis. The size of the dot represents the relative number of consistent DE genes, which for selected upstream regulator is 131.* **b) Volcano plot:** *There are **131** DE genes that are targets of Dihydrotestosterone consistent with the hypothesis that Dihydrotestosterone is present (overly abundant) The target genes are represented in terms of their measured expression change (x-axis) and the significance of the change (y-axis). The significance is represented in terms of the negative log (base 10) of the p-value, so that more significant genes are plotted higher on the y-axis.*

#### cylindrospermopsin

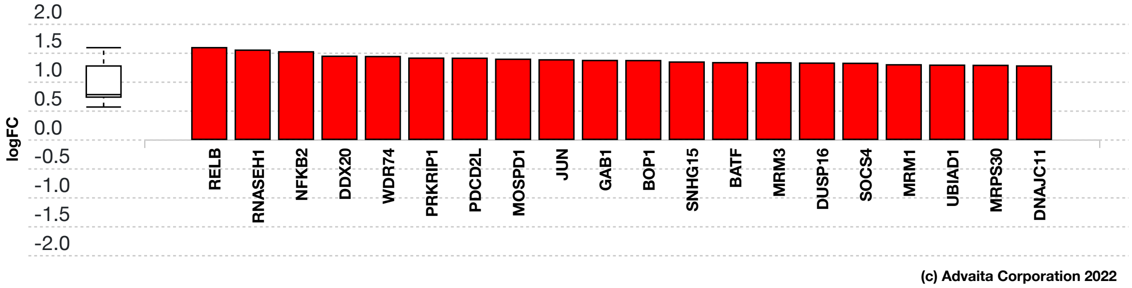


**Fig. 6.2.9: Consistent DE target genes measured expression bar plot:** *All the consistent differentially expressed genes that are targeted by cylindrospermopsin are ranked based on their absolute value of log fold change. The plot is limited to the top 20 genes out of a total of 77 consistent differentially expressed target genes. Upregulated genes are shown in red, downregulated genes are shown in blue. The box and whisker plot on the left summarizes the distribution of all the consistent differentially expressed genes targeted by this upstream regulator. The box shows the 1st quartile, the median and the 3rd quartile, while any outliers are represented by circles.*

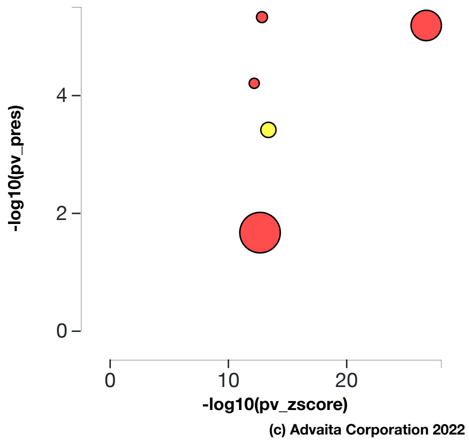


a)

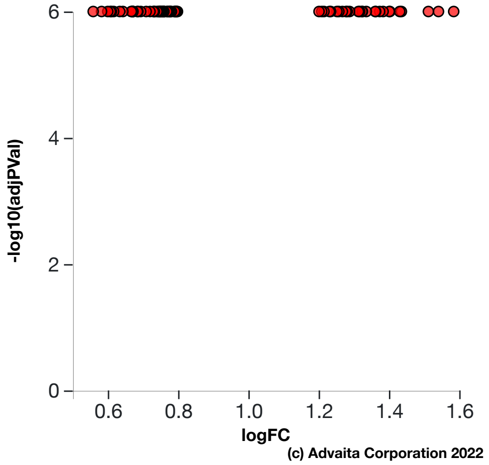


b)

**Fig. 6.2.10: a) Present (overly abundant) p-value vs zscore p-value:** *The significance of cylindrospermopsin is plotted on two axes, with negative log of $P_z$ on x-axis and negative log of $P_{pres}$ on y-axis. The size of the dot represents the relative number of consistent DE genes, which for selected upstream regulator is 77.* **b) Volcano plot:** *There are **77** DE genes that are targets of cylindrospermopsin consistent with the hypothesis that cylindrospermopsin is present (overly abundant) The target genes are represented in terms of their measured expression change (x-axis) and the significance of the change (y-axis). The significance is represented in terms of the negative log (base 10) of the p-value, so that more significant genes are plotted higher on the y-axis.*

#### Sodium Selenite

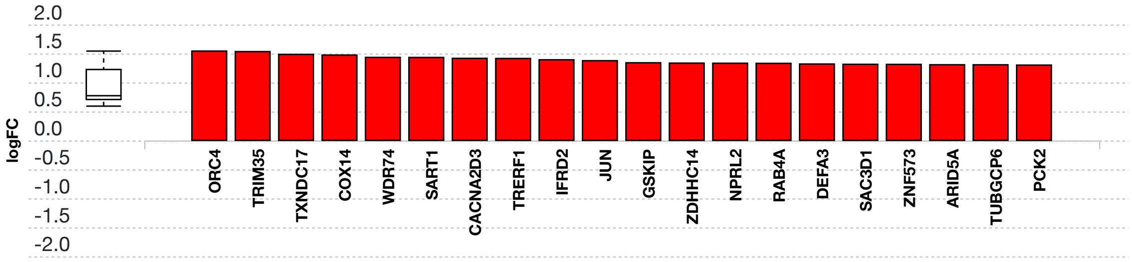


**Fig. 6.2.11: Consistent DE target genes measured expression bar plot:** *All the consistent differentially expressed genes that are targeted by Sodium Selenite are ranked based on their absolute value of log fold change. The plot is limited to the top 20 genes out of a total of 167 consistent differentially expressed target genes. Upregulated genes are shown in red, downregulated genes are shown in blue. The box and whisker plot on the left summarizes the distribution of all the consistent differentially expressed genes targeted by this upstream regulator. The box shows the 1st quartile, the median and the 3rd quartile, while any outliers are represented by circles.*

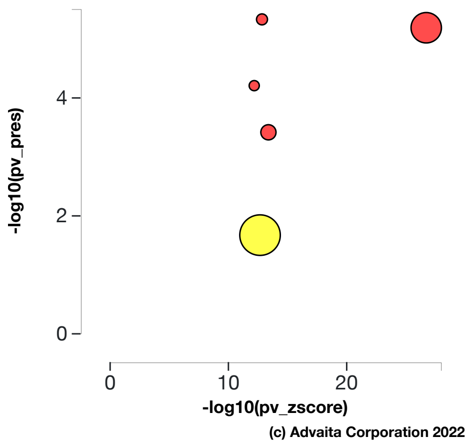


a)

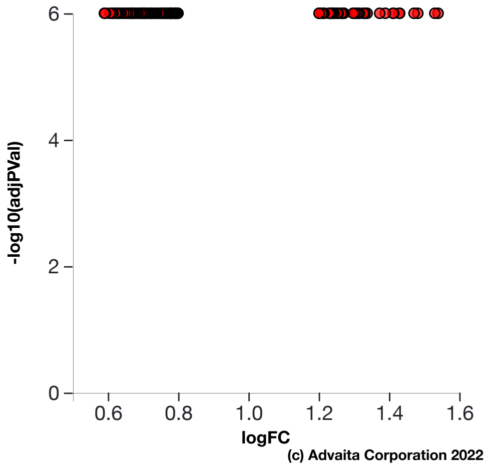


b)

**Fig. 6.2.12: a) Present (overly abundant) p-value vs zscore p-value:** *The significance of Sodium Selenite is plotted on two axes, with negative log of $P_z$ on x-axis and negative log of $P_{pres}$ on y-axis. The size of the dot represents the relative number of consistent DE genes, which for selected upstream regulator is 167.* **b) Volcano plot:** *There are **167** DE genes that are targets of Sodium Selenite consistent with the hypothesis that Sodium Selenite is present (overly abundant) The target genes are represented in terms of their measured expression change (x-axis) and the significance of the change (y-axis). The significance is represented in terms of the negative log (base 10) of the p-value, so that more significant genes are plotted higher on the y-axis.*

### 6.3. Results: upstream CDTs predicted as absent (or insufficient)

| CDT (u) | DTI(u) | DT(u) | p-value | p-value (FDR) | p-value (Bonferroni) |
|---|---|---|---|---|---|
| Doxorubicin | 932 | 1131 | **5.626e-23** | **3.355e-20** | **1.243e-19** |
| Ivermectin | 833 | 848 | **5.626e-23** | **3.355e-20** | **1.243e-19** |
| dicrotophos | 499 | 696 | **5.626e-23** | **3.355e-20** | **1.243e-19** |
| 3-((6-(2-methoxyphenyl)pyrimidin-4-yl)amino)phenyl)methane sulfonamide | 162 | 167 | **6.075e-23** | **3.355e-20** | **1.342e-19** |
| 7,8-Dihydro-7,8-dihydroxybenzo(a)pyrene 9,10-oxide | 440 | 563 | **7.519e-19** | **3.322e-16** | **1.661e-15** |

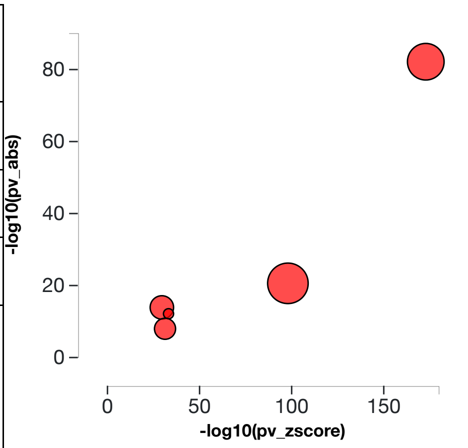


**Table 6.3.1: Top upstream CDTs predicted as absent (or insufficient).** *For each upstream CDT u, the table shows the number of DE targets supporting the hypothesis that the CDT is absent DTI(u) the total number of DE genes downstream of u DT(u), the combined raw p-value, and the p-value corrected for multiple comparisons.* **Fig. 6.3.1: A two-way plot showing the top five upstream CDTs predicted as absent (or insufficient).** *Dots representing upstream CDTs are positioned using $P_{zscore}$ on the horizontal axis, and using $P_{abs}$ on the vertical axis. $P_{abs}$ is the p-value based on the number of DE targets consistent with the typeof the incoming signal and with the selected hypothesis type. Upstream CDTs with a significant combined p-value are shown in red. The size of each dot represents the relative number of consistent DE genes for that CDT.*

#### Doxorubicin

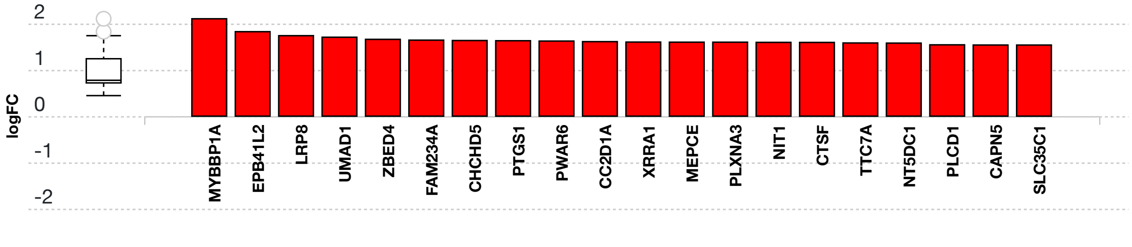


**Fig. 6.3.13: Consistent DE target genes measured expression bar plot:** *All the consistent differentially expressed genes that are targeted by Doxorubicin are ranked based on their absolute value of log fold change. The plot is limited to the top 20 genes out of a total of 932 consistent differentially expressed target genes. Upregulated genes are shown in red, downregulated genes are shown in blue. The box and whisker plot on the left summarizes the distribution of all the consistent differentially expressed genes targeted by this upstream regulator. The box shows the 1st quartile, the median and the 3rd quartile, while any outliers are represented by circles.*

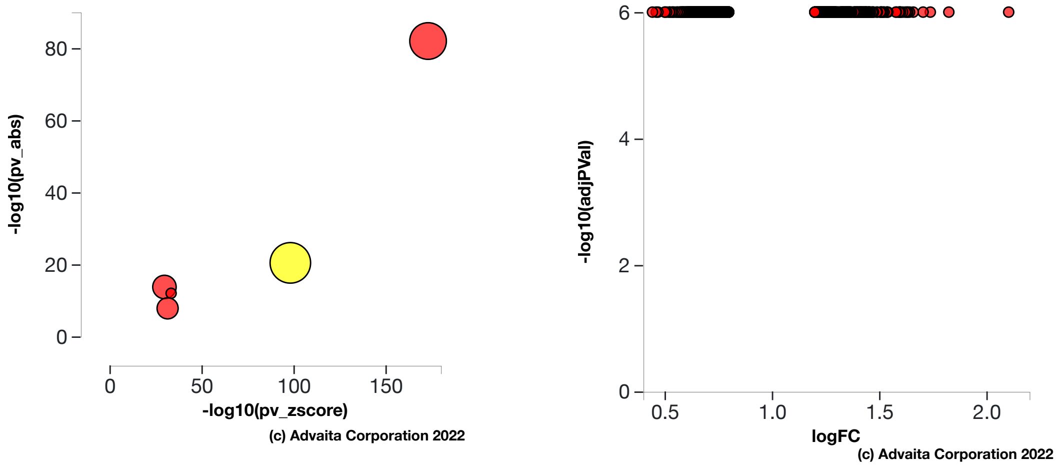


a) b)

**Fig. 6.3.14: a) Absent (or insufficient) p-value vs zscore p-value:** *The significance of Doxorubicin is plotted on two axes, with negative log of $P_z$ on x-axis and negative log of $P_{abs}$ on y-axis. The size of the dot represents the relative number of consistent DE genes, which for selected upstream regulator is 932.* **b) Volcano plot:** *There are **932** DE genes that are targets of Doxorubicin consistent with the hypothesis that Doxorubicin is absent (or insufficient) The target genes are represented in terms of their measured expression change (x-axis) and the significance of the change (y-axis). The significance is represented in terms of the negative log (base 10) of the p-value, so that more significant genes are plotted higher on the y-axis.*

#### Ivermectin

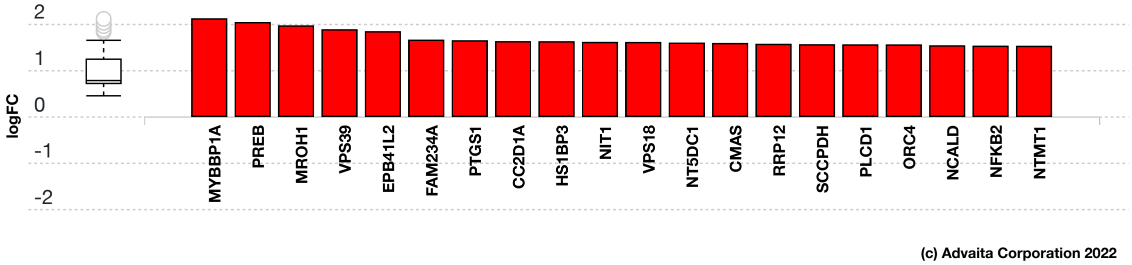


**Fig. 6.3.15: Consistent DE target genes measured expression bar plot:** *All the consistent differentially expressed genes that are targeted by Ivermectin are ranked based on their absolute value of log fold change. The plot is limited to the top 20 genes out of a total of 833 consistent differentially expressed target genes. Upregulated genes are shown in red, downregulated genes are shown in blue. The box and whisker plot on the left summarizes the distribution of all the consistent differentially expressed genes targeted by this upstream regulator. The box shows the 1st quartile, the median and the 3rd quartile, while any outliers are represented by circles.*

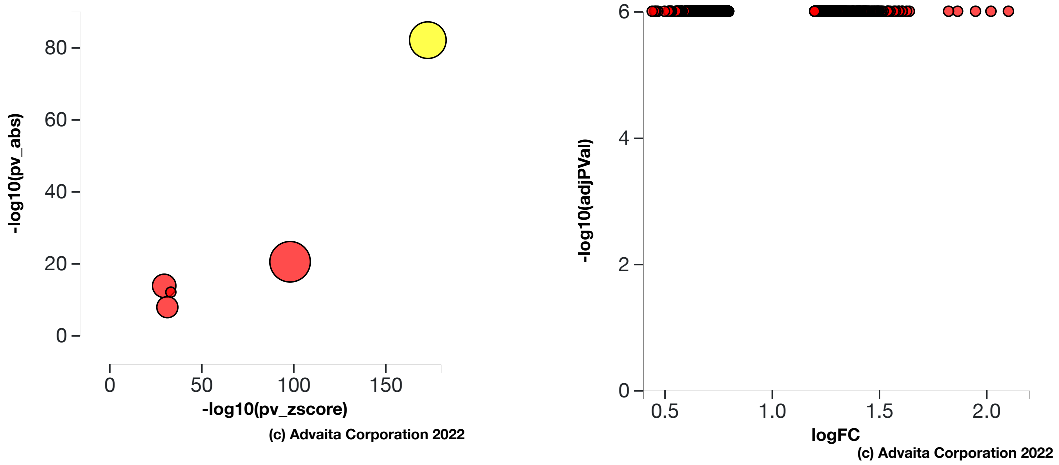


a) b)

**Fig. 6.3.16: a) Absent (or insufficient) p-value vs zscore p-value:** *The significance of Ivermectin is plotted on two axes, with negative log of $P_z$ on x-axis and negative log of $P_{abs}$ on y-axis. The size of the dot represents the relative number of consistent DE genes, which for selected upstream regulator is 833.* **b) Volcano plot:** *There are **833** DE genes that are targets of Ivermectin consistent with the hypothesis that Ivermectin is absent (or insufficient) The target genes are represented in terms of their measured expression change (x-axis) and the significance of the change (y-axis). The significance is represented in terms of the negative log (base 10) of the p-value, so that more significant genes are plotted higher on the y-axis.*

#### dicrotophos

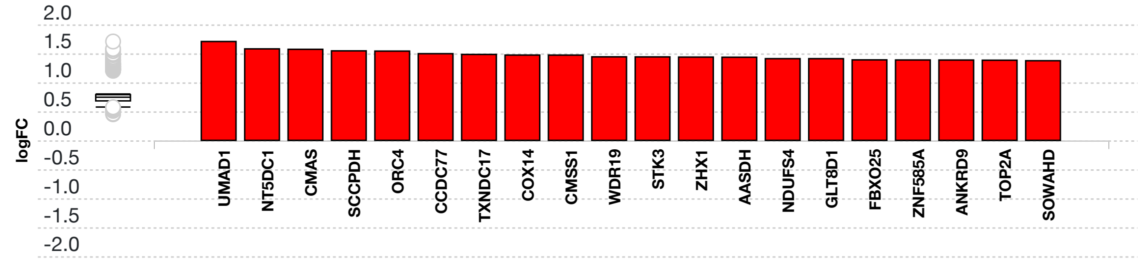


**Fig. 6.3.17: Consistent DE target genes measured expression bar plot:** *All the consistent differentially expressed genes that are targeted by dicrotophos are ranked based on their absolute value of log fold change. The plot is limited to the top 20 genes out of a total of 499 consistent differentially expressed target genes. Upregulated genes are shown in red, downregulated genes are shown in blue. The box and whisker plot on the left summarizes the distribution of all the consistent differentially expressed genes targeted by this upstream regulator. The box shows the 1st quartile, the median and the 3rd quartile, while any outliers are represented by circles.*

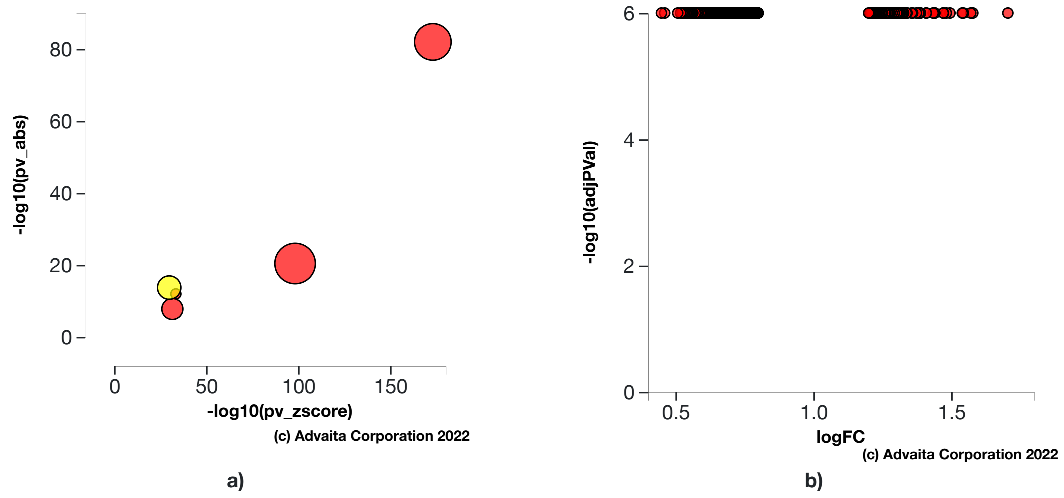


**Fig. 6.3.18: a) Absent (or insufficient) p-value vs zscore p-value:** *The significance of dicrotophos is plotted on two axes, with negative log of $P_z$ on x-axis and negative log of $P_{abs}$ on y-axis. The size of the dot represents the relative number of consistent DE genes, which for selected upstream regulator is 499.* **b) Volcano plot:** *There are* ***499*** *DE genes that are targets of dicrotophos consistent with the hypothesis that dicrotophos is absent (or insufficient) The target genes are represented in terms of their measured expression change (x-axis) and the significance of the change (y-axis). The significance is represented in terms of the negative log (base 10) of the p-value, so that more significant genes are plotted higher on the y-axis.*

#### 3-((6-(2-methoxyphenyl)pyrimidin-4-yl)amino)phenyl)methane sulfonamide

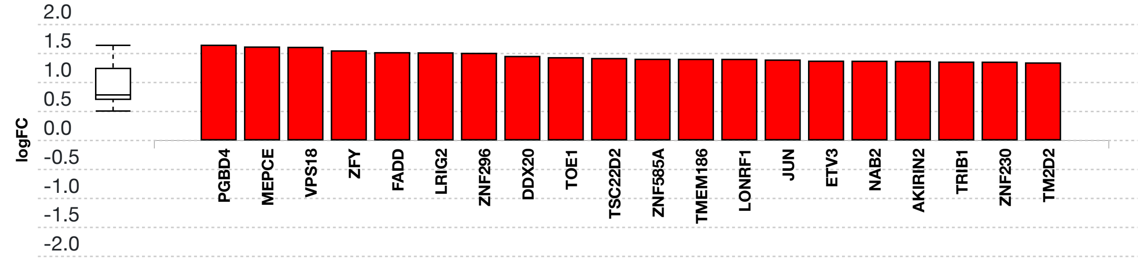


**Fig. 6.3.19: Consistent DE target genes measured expression bar plot:** *All the consistent differentially expressed genes that are targeted by 3-((6-(2-methoxyphenyl)pyrimidin-4-yl)amino)phenyl)methane sulfonamide are ranked based on their absolute value of log fold change. The plot is limited to the top 20 genes out of a total of 162 consistent differentially expressed target genes. Upregulated genes are shown in red, downregulated genes are shown in blue. The box and whisker plot on the left summarizes the distribution of all the consistent differentially expressed genes targeted by this upstream regulator. The box shows the 1st quartile, the median and the 3rd quartile, while any outliers are represented by circles.*

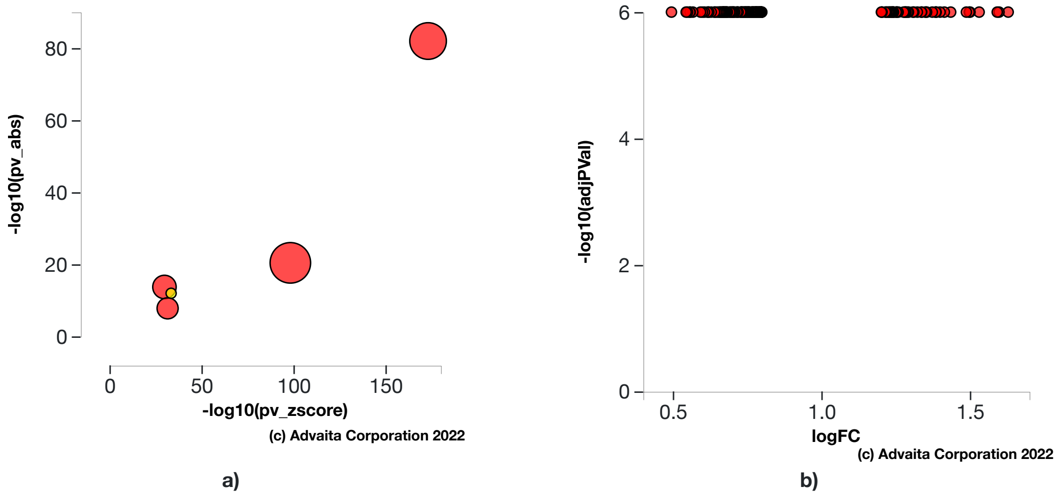


**Fig. 6.3.20: a) Absent (or insufficient) p-value vs zscore p-value:** *The significance of 3-((6-(2-methoxyphenyl)pyrimidin-4-yl)amino)phenyl)methane sulfonamide is plotted on two axes, with negative log of $P_z$ on x-axis and negative log of $P_{abs}$ on y-axis. The size of the dot represents the relative number of consistent DE genes, which for selected upstream regulator is 162.* **b) Volcano plot:** *There are* ***162*** *DE genes that are targets of 3-((6-(2-methoxyphenyl)pyrimidin-4-yl)amino)phenyl)methane sulfonamide consistent with the hypothesis that 3-((6-(2-methoxyphenyl)pyrimidin-4-yl)amino)phenyl)methane sulfonamide is absent (or insufficient) The target genes are represented in terms of their measured expression change (x-axis) and the significance of the change (y-axis). The significance is represented in terms of the negative log (base 10) of the p-value, so that more significant genes are plotted higher on the y-axis.*

#### 7,8-Dihydro-7,8-dihydroxybenzo(a)pyrene 9,10-oxide

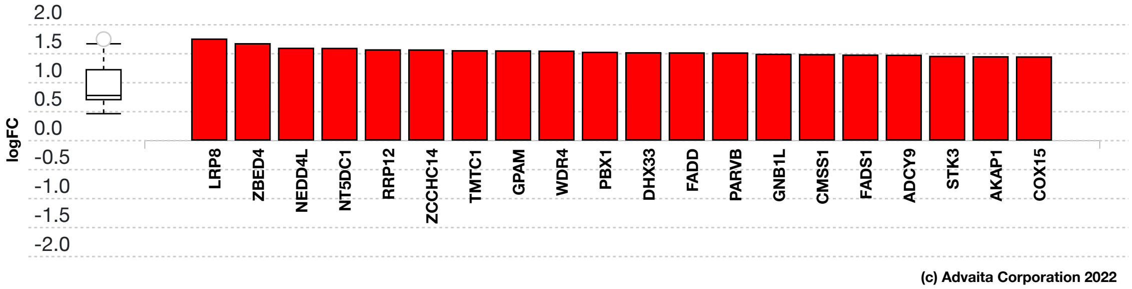


**Fig. 6.3.21: Consistent DE target genes measured expression bar plot:** *All the consistent differentially expressed genes that are targeted by 7,8-Dihydro-7,8-dihydroxybenzo(a)pyrene 9,10-oxide are ranked based on their absolute value of log fold change. The plot is limited to the top 20 genes out of a total of 440 consistent differentially expressed target genes. Upregulated genes are shown in red, downregulated genes are shown in blue. The box and whisker plot on the left summarizes the distribution of all the consistent differentially expressed genes targeted by this upstream regulator. The box shows the 1st quartile, the median and the 3rd quartile, while any outliers are represented by circles.*

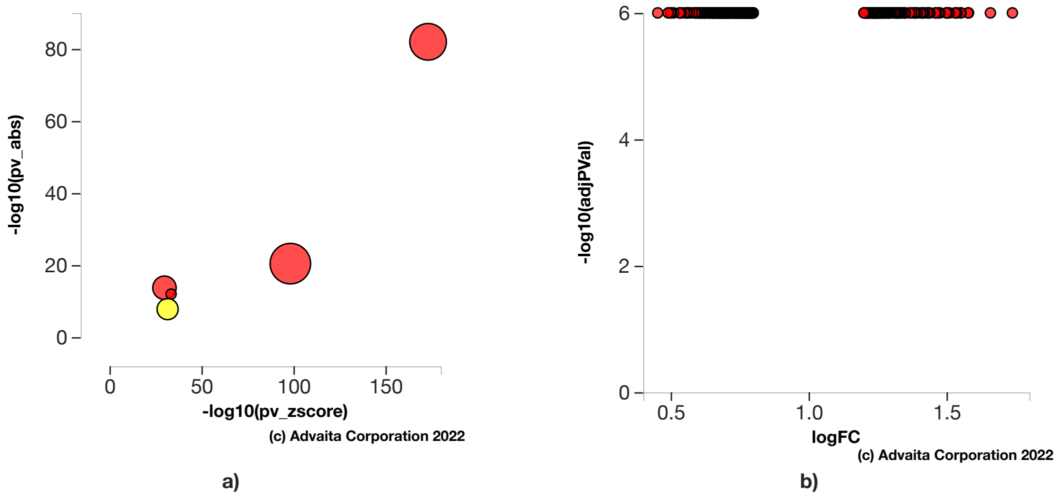


**Fig. 6.3.22: a) Absent (or insufficient) p-value vs zscore p-value:** *The significance of 7,8-Dihydro-7,8-dihydroxybenzo(a)pyrene 9,10-oxide is plotted on two axes, with negative log of $P_z$ on x-axis and negative log of $P_{abs}$ on y-axis. The size of the dot represents the relative number of consistent DE genes, which for selected upstream regulator is 440.* **b) Volcano plot:** *There are* ***440*** *DE genes that are targets of 7,8-Dihydro-7,8-dihydroxybenzo(a)pyrene 9,10-oxide consistent with the hypothesis that 7,8-Dihydro-7,8-dihydroxybenzo(a)pyrene 9,10-oxide is absent (or insufficient) The target genes are represented in terms of their measured expression change (x-axis) and the significance of the change (y-axis). The significance is represented in terms of the negative log (base 10) of the p-value, so that more significant genes are plotted higher on the y-axis.*

### 7. Disease Analysis

### 7.1. Methods

For each disease, the number of differentially expressed (DE) genes annotated to a disease term is compared to the number of DE genes expected just by chance. iPathwayGuide uses an over-representation approach to compute the statistical significance of observing at least the given number of DE genes. The p-value is computed using the hypergeometric distribution as described for pORA in the Pathway Analysis section. This p-value is corrected for multiple comparisons using FDR and Bonferroni.

### 7.2. Results

**Table 7.2.1: Top identified diseases**

| Disease Name | p-value | p-value (FDR) | p-value (Bonferroni) |
|---|---|---|---|
| Congenital disorders of glycosylation type I | 5.272e-8 | 1.763e-5 | 2.673e-5 |
| Autosomal recessive mental retardation | 6.954e-8 | 1.763e-5 | 3.526e-5 |
| Joubert syndrome | 9.091e-7 | 1.536e-4 | 4.609e-4 |
| Pontocerebellar hypoplasia | 2.861e-6 | 2.901e-4 | 0.001 |
| Cytochrome c oxidase (COX) deficiency; Mitochondrial complex IV deficiency (MT-C4D) | 2.861e-6 | 2.901e-4 | 0.001 |

#### Congenital disorders of glycosylation type I (H00118)

Congenital disorders of glycosylation (CDG) are a group of disorders caused by defects in various genes for N-glycan biosynthesis. CDG type I is defined by mutations in genes encoding enzymes which involves disrupted synthesis of the lipid linked oligosaccharide precursor and its transfer to polypeptide chain of protein, affecting N-glycan assembly in cytosol and endoplasmic reticulum. An increasing number of disorders have been discovered, with many subtypes identified. PMM2-CDG is the most common form, with over 800 patients diagnosed mostly in Europe. Almost all type present in infancy. These diseases demonstrate a broad range of clinical manifestation, associated with developmental delay, psychomotor retardation, hypotonia, seizures, hepatomegaly, microcephaly, and pericardial effusion. In this experiment, the algorithm identified **11** differentially expressed genes out of **29** genes associated with the disease.

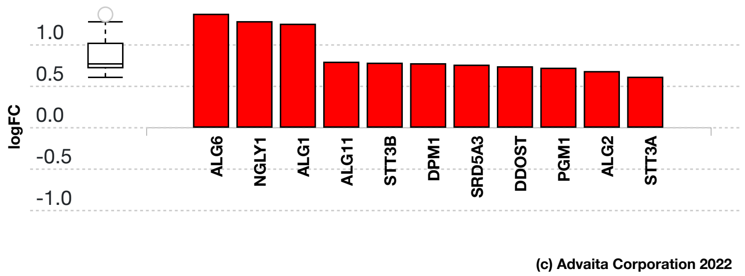


**Fig. 7.2.1: Gene measured expression bar plot:** *All the differentially expressed genes that are annotated to Congenital disorders of glycosylation type I are ranked based on their absolute value of log fold change. Upregulated genes are shown in red, downregulated genes are shown in blue. The box plot on the left summarizes the distribution of all the differentially expressed genes that are annotated to this disease. The box represents the 1st quartile, the median and the 3rd quartile, while the outliers are represented by circles.*

#### Autosomal recessive mental retardation (H00768)

Mental retardation (MR) is a neurodevelopmental disorder characterized by low intelligence quotient (IQ) and deficits in adaptive behaviors. Although X-linked MR has been extensively studied, and over 80 causal genes have been cloned, little is known about the genetic basis of autosomal recessive mental retardation (MRT). To date, several genes have been identified. These genes have a variety of functions and participate in multiple biochemical pathways. In addition, there are several known disease loci for which genes have not yet been identified. In this experiment, the algorithm identified **14** differentially expressed genes out of **50** genes associated with the disease.

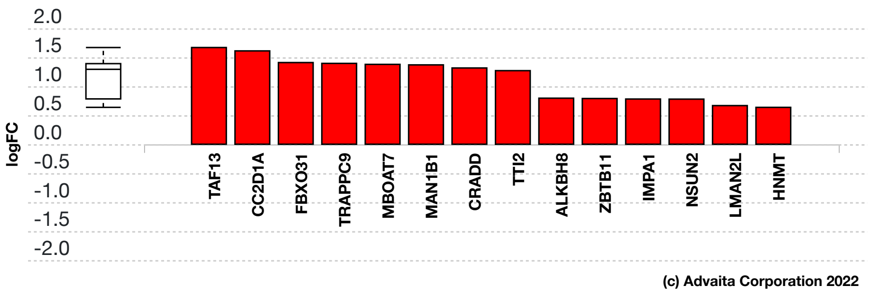


**Fig. 7.2.2: Gene measured expression bar plot:** *All the differentially expressed genes that are annotated to Autosomal recessive mental retardation are ranked based on their absolute value of log fold change. Upregulated genes are shown in red, downregulated genes are shown in blue. The box plot on the left summarizes the distribution of all the differentially expressed genes that are annotated to this disease. The box represents the 1st quartile, the median and the 3rd quartile, while the outliers are represented by circles.*

#### Joubert syndrome (H00530)

Joubert syndrome and related disorders are a group of multiple congenital anomaly syndromes characterized by 'molar tooth sign', a specific midbrain-hindbrain malformation seen in brain images. Joubert syndrome is associated with retinal dystrophy, nephronophthisis, liver fibrosis and polydactyly. Most of the causative genes encode cilium-related proteins. In this experiment, the algorithm identified **11** differentially expressed genes out of **37** genes associated with the disease.

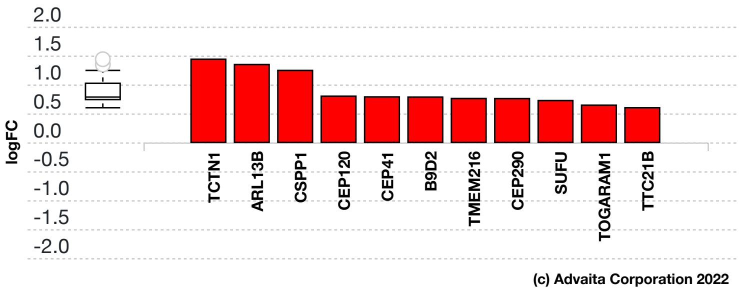


**Fig. 7.2.3: Gene measured expression bar plot:** *All the differentially expressed genes that are annotated to Joubert syndrome are ranked based on their absolute value of log fold change. Upregulated genes are shown in red, downregulated genes are shown in blue. The box plot on the left summarizes the distribution of all the differentially expressed genes that are annotated to this disease. The box represents the 1st quartile, the median and the 3rd quartile, while the outliers are represented by circles.*

#### Pontocerebellar hypoplasia (H00897)

Pontocerebellar hypoplasia (PCH) is a group of inherited progressive neurodegenerative disorders with prenatal onset. Up to now ten different subtypes have been reported. All subtypes share common characteristics, including hypoplasia/atrophy of cerebellum and pons, progressive microcephaly, and variable cerebral involvement. Mutations in three tRNA splicing endonuclease subunit genes were found to be responsible for PCH2, PCH4 and PCH5. Mutations in the nuclear encoded mitochondrial arginyl- tRNA synthetase gene underlie PCH6. PCH1 is caused by homozygous mutation in the VRK1 gene. In this experiment, the algorithm identified **7** differentially expressed genes out of **15** genes associated with the disease.

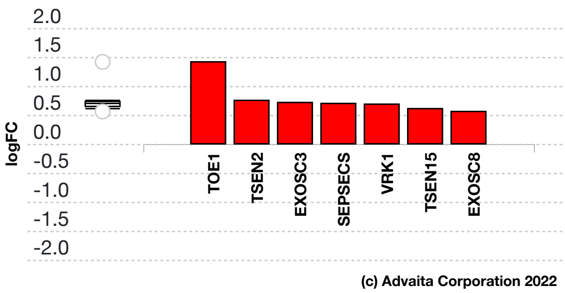


**Fig. 7.2.4: Gene measured expression bar plot:** *All the differentially expressed genes that are annotated to Pontocerebellar hypoplasia are ranked based on their absolute value of log fold change. Upregulated genes are shown in red, downregulated genes are shown in blue. The box plot on the left summarizes the distribution of all the differentially expressed genes that are annotated to this disease. The box represents the 1st quartile, the median and the 3rd quartile, while the outliers are represented by circles.*

##### Cytochrome c oxidase (COX) deficiency; Mitochondrial complex IV deficiency (MT-C4D) (H01368)

Cytochrome c oxidase (COX) deficiency is a mitochondrial disease that is caused by the lack of the COX. Cytochrome c oxidase (COX) is the terminal enzyme of the mitochondrial respiratory chain (complex IV). Since COX is encoded by nuclear and mitochondrial genes, COX deficiency can be inherited in either an autosomal recessive or a maternal pattern. Patients can present with a number of different clinical phenotypes, including Leigh syndrome, Fatal infantile cardioencephalomyopathy, and Leber hereditary optic neuropathy. In this experiment, the algorithm identified **7** differentially expressed genes out of **15** genes associated with the disease.

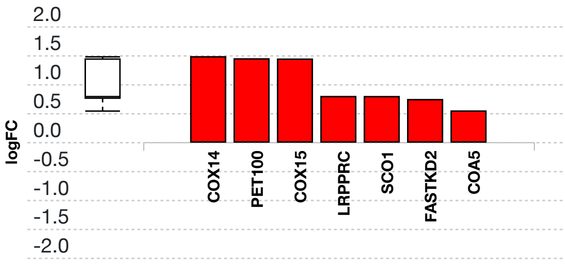


**Fig. 7.2.5: Gene measured expression bar plot:** *All the differentially expressed genes that are annotated to Cytochrome c oxidase (COX) deficiency; Mitochondrial complex IV deficiency (MT-C4D) are ranked based on their absolute value of log fold change. Upregulated genes are shown in red, downregulated genes are shown in blue. The box plot on the left summarizes the distribution of all the differentially expressed genes that are annotated to this disease. The box represents the 1st quartile, the median and the 3rd quartile, while the outliers are represented by circles.*
